# Global Trends in Cardiovascular Mortality and Major Cardiometabolic Risk Factors, 2000–2023: A Longitudinal Ecological Analysis of 204 Countries and Territories

**DOI:** 10.64898/2026.09.14.26363013

**Authors:** Khalid Mohammed Al-Dhayani

## Abstract

**Background and Aims:** Cardiovascular disease (CVD) remains the leading cause of global mortality. We evaluated 24-year global trends in age-standardized CVD mortality across 204 countries and territories (2000–2023) and decomposed the independent longitudinal associations of major cardiometabolic risk factor pathways.

**Methods:** The study analyzed a balanced panel of 4,896 country-year observations using Global Burden of Disease 2023 and World Bank data. Two-way fixed-effects panel regression models adjusting for country fixed effects, year fixed effects, GDP per capita, and urbanization were specified using Driscoll–Kraay robust standard errors. Hierarchical path mediation and extended 1- to 10-year lag models evaluated risk pathways and disease latency.

**Results:** Global age-standardized CVD mortality declined from 340.2 ± 156.4 to 258.4 ± 132.1 deaths per 100,000 population (2000–2023), but annual declines stagnated post-2019 (−1.31% to −0.03% per year). High SBP (adjusted beta = 1.129), high LDL cholesterol (beta = 0.622), smoking (beta = 0.452), and high fasting glucose (beta = 0.410) were independently associated with CVD mortality (P < 0.001 for all). High SBP reduction accounted for 70.91% of the global mortality decline. High BMI was not directly associated with mortality (beta = 0.049, P = 0.421); mediation analysis proved that 99.20% of its population impact is mediated through downstream metabolic intermediaries.

**Conclusions:** Elevated SBP and cumulative LDL cholesterol are the primary drivers of global cardiovascular mortality. Obesity acts as an upstream driver operating almost entirely through downstream metabolic intermediaries. Sustained progress requires prioritizing universal blood pressure control, lipid lowering, and multi-sectoral obesity prevention.

## 1. INTRODUCTION

Cardiovascular diseases (CVDs) remain the leading cause of premature mortality and healthcare burden worldwide, accounting for approximately one-third of all deaths worldwide, despite major advances in preventive cardiology, therapeutic pharmacotherapy, and invasive interventions (1,2,5). Although age-standardized CVD mortality rates have declined in many regions over recent decades owing to expanded hypertension control, lipid-lowering therapy, smoking cessation campaigns, and improved acute coronary care, the absolute global number of cardiovascular deaths continues to rise as a result of population growth and demographic aging (1–3,5). Consequently, CVD remains a formidable global public health challenge, with nearly three-quarters of all cardiovascular deaths occurring in low- and middle-income countries (LMICs), where healthcare resources, diagnostic infrastructure, and preventive services remain severely constrained (5–8).

Metabolic and behavioral risk factors—specifically elevated systolic blood pressure (SBP), elevated low-density lipoprotein (LDL) cholesterol, hyperglycemia, obesity, and tobacco smoking—are the primary modifiable drivers of cardiovascular morbidity and mortality worldwide (1,5,7,12). Among these, hypertension remains the single largest preventable contributor to cardiovascular death globally (7,24,25), whereas the increasing global prevalence of obesity and type 2 diabetes threatens to erode gains achieved through traditional cardiovascular prevention (9,19,20,23). However, evaluating the relative population-level contributions of these individual risk factors is complicated by their frequent clustering within metabolic syndrome, creating substantial collinearity among blood pressure, atherogenic lipids, impaired glucose tolerance, and excess adiposity (21–23).

Previous studies, particularly those derived from the Global Burden of Disease (GBD) framework, have comprehensively described descriptive trends in cardiovascular mortality and individual risk factor prevalence (1,2,5). Nevertheless, most existing studies have relied on descriptive cross-sectional estimates or unadjusted attributable burden metrics (population attributable fractions) rather than simultaneously evaluating the adjusted longitudinal associations of multiple cardiometabolic risk pathways within a unified econometric panel model. Furthermore, cross-sectional analyses often fail to control for unobserved time-invariant country characteristics (such as baseline institutional, geographic, or health-system differences) and common global secular trends (such as worldwide therapeutic advances or global economic disruptions) (37,38).

To address these methodological limitations, we conducted a comprehensive global longitudinal ecological panel analysis encompassing 204 countries and territories over a 24-year period (2000–2023), yielding 4,896 country-year observations. By utilizing a two-way fixed-effects (TWFE) panel regression design incorporating Driscoll–Kraay robust standard errors (38), this study aimed to (1) evaluate 24-year secular trends in age-standardized CVD mortality and major risk factor-attributable mortality globally and across World Bank income groups and World Health Organization (WHO) regions; (2) quantify the independent population-level associations of SBP, LDL cholesterol, fasting plasma glucose, BMI, and smoking with CVD mortality after simultaneous multivariable adjustment; and (3) examine the stability and potential heterogeneity of these associations across alternative model specifications, temporal lag structures, nonlinear functional forms, and macroeconomic settings.

## 2. METHODS

### 2.1 Study Design and Analytical Sample

This study conducted a longitudinal ecological panel study to examine temporal patterns in age-standardized CVD mortality and their population-level associations with major cardiometabolic and behavioral risk factors across 204 countries and territories from 2000 through 2023. The unit of analysis was the country-year observation, with each observation representing one country in one calendar year. The final analytical dataset comprises a complete, balanced longitudinal panel of 4,896 country-year observations (204 countries and territories observed annually over 24 consecutive years).

The study was designed to evaluate population-level longitudinal associations rather than individual-level causal relationships. Accordingly, estimates represent macrolevel relationships between country-level changes in risk factor-attributable mortality and age-standardized CVD mortality and should not be interpreted as individual-level effects.

### 2.2 Data sources and variable definitions

Annual country-level data on CVD mortality and risk factor-attributable mortality were obtained from the Institute for Health Metrics and Evaluation (IHME) GBD 2023 study database. The GBD framework provides annual, age-standardized estimates of cause-specific mortality and mortality attributable to established cardiovascular risk factors via standardized comparative risk assessment methods intended to facilitate comparisons across countries and over time.

Annual national socioeconomic indicators, including gross domestic product (GDP) per capita adjusted for purchasing power parity (PPP, expressed in constant international US dollars) and the percentage of the total population residing in urban areas, were obtained from the World Bank Open Data repository. These variables were incorporated as time-varying country-level covariates to account for socioeconomic development and urbanization.

Data from the GBD and World Bank sources were harmonized by country and calendar year. Countries and territories were eligible for inclusion when complete annual information was available for all variables required for the primary analysis throughout 2000–2023. The resulting dataset included 204 countries and territories and 4,896 country-year observations.

### 2.3 Outcome and Exposure Variables

The primary outcome variable was the age-standardized CVD mortality rate, expressed in standard SI units as deaths per 100,000 people. Age-standardized mortality rates were used to improve comparability between countries and across years by accounting for differences and changes in population age structure over time. The primary exposures were age-standardized CVD mortality rates (expressed as deaths per 100,000 people) attributable to five major risk factors:

1. High SBP
2. High LDL cholesterol
3. High fasting plasma glucose
4. High BMI
5. Tobacco smoking

All risk factor measures were derived from the GBD 2023 database and analyzed as continuous country-level measures. These variables represent population-level mortality attributable to the corresponding risk factors and should not be interpreted as individual exposure prevalence or individual-level risk estimates.

The principal time-varying covariates were GDP per capita (PPP international dollars) and the urban population percentage. Country fixed effects were included to account for unobserved characteristics that remain constant within countries over time, including persistent geographic, demographic, cultural, institutional, and health-system characteristics. Calendar-year fixed effects were included to account for secular trends and common temporal shocks occurring across countries in a given year.

### 2.4 Statistical analysis

#### Descriptive and temporal analyses

Continuous variables were summarized using means and standard deviations (SDs). Temporal patterns in age-standardized CVD mortality and risk factor-attributable mortality were examined over the 2000–2023 study period. Annual percentage changes (APCs) were calculated to characterize year-over-year temporal changes in mortality measures.

Pearson correlation coefficients (r) were calculated as descriptive measures of the univariable associations between CVD mortality and individual risk factor measures. These analyses were considered exploratory and were not used to infer independent associations after adjustment for country and year fixed effects.

#### Primary two-way fixed-effects model

The primary analysis used the TWFE panel regression model to estimate the independent longitudinal associations between risk factor-attributable mortality and age-standardized CVD mortality.

The model was specified as follows:

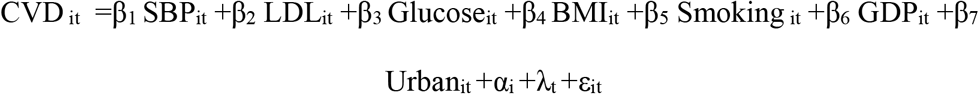

where CVD_it_ denotes the age-standardized CVD mortality rate for country i in year ***t***; SBP_it_, LDL_it_, Glucose_it_, BMI_it_, and Smoking_it_ denote mortality attributable to high systolic blood pressure, high LDL cholesterol, high fasting plasma glucose, high BMI, and smoking, respectively; GDP_it_ represents GDP per capita; and Urban_it_ represents the percentage of the population living in urban areas. The term α_i_ represents country fixed effects, λ_t_ represents calendar-year fixed effects, and ɛ_it_ represents the idiosyncratic error term.

#### Robust inference and model diagnostics

Preliminary diagnostic assessment indicated substantial violations of conventional independent-error assumptions. The Durbin–Watson statistic was 0.0989, indicating strong positive serial correlation, whereas the Breusch–Pagan test indicated significant heteroskedasticity (LM statistic = 618.18; P < 0.001). Residual normality was also rejected by the Lilliefors test (P = 0.001).

Because the panel data exhibited heteroskedasticity, serial correlation, and dependence across countries, inference for the primary TWFE model was based on Driscoll–Kraay robust standard errors. This estimator provides covariance estimates robust to heteroskedasticity, serial correlation, and broad forms of cross-sectional spatial dependence in panel data. Thus, regression coefficients (beta), standard errors (SE), 95% confidence intervals (CIs), and P values reported for the primary model were calculated via the Driscoll–Kraay covariance estimator. The nonnormality of residuals was not treated as a basis for excluding the model because inference was based on robust panel covariance estimation, and the analysis involved a large number of country–year observations.

#### Comparative Model Specifications

To assess the contribution of year fixed effects, estimates from a country fixed-effects model (controlling for time-invariant country characteristics without common year fixed effects) were compared with those from the primary TWFE model (incorporating both country and calendar-year fixed effects). The stability of coefficient estimates across these specifications was evaluated as part of sensitivity testing.

#### Sensitivity analyses

Several prespecified sensitivity analyses were conducted to evaluate the robustness of the primary findings:

1. COVID-19 sensitivity analysis: Because the COVID-19 pandemic has substantially affected mortality patterns, healthcare utilization, and cause-of-death attribution, a sensitivity analysis excluded observations from 2020–2021. The model was re-estimated via the remaining country-year observations to determine whether the principal associations persisted independently of the pandemic period.
2. Lagged Analyses: To examine temporal ordering and reduce concerns regarding contemporaneous associations, 1-year, 3-year, and 5-year lagged analyses were conducted. Risk factor measures were lagged relative to CVD mortality such that risk factor-attributable mortality in year (t-k) was related to CVD mortality in year (t). The GDP per capita and urban population were retained as covariates.

#### Nonlinearity and heterogeneity analyses

Potential departures from linearity were examined by extending the regression model to include quadratic terms (Risk squared) for high SBP, high LDL cholesterol, high fasting plasma glucose, high BMI, and smoking. The significance and direction of the corresponding squared terms were evaluated to determine whether the statistical evidence supported nonlinear relationships.

Potential heterogeneity in associations was evaluated according to the World Bank income group (low, lower-middle, upper-middle, and high income) and WHO region (AFRO, AMRO, EMRO, EURO, SEARO, and WPRO). Interaction terms were introduced between each principal risk factor and country income-group categories and WHO regions. Interaction coefficients were interpreted in conjunction with 95% confidence intervals and joint Wald test statistics.

#### Multicollinearity, influence, and missing data

Potential multicollinearity among explanatory variables was evaluated via variance inflation factors (VIFs) and pairwise correlations. Influence diagnostics were conducted via Cook’s distance to identify country-year observations exceeding the prespecified influence threshold (4/n = 0.00082). Leave-one-country-out (LOCO) sensitivity analyses were conducted to evaluate model stability when highly influential countries were excluded.

The primary analysis was based on the complete balanced panel of 4,896 country-year observations. Because the analysis used aggregated national-level data, no individual-level missingness or participant-level loss to follow-up was applicable. All the statistical tests were two-sided, and P < 0.05 was considered statistically significant.

#### Computational Software, Reproducibility, and AI Tool Usage

All the statistical analyses were performed via Python version 3.12.x (Python Software Foundation). Data manipulation and variable construction were conducted via pandas version 2.2.2 and NumPy version 2.0.2. Panel regression models were estimated via the PanelOLS estimator in linearmodels version 7.0 with Driscoll–Kraay robust covariance estimation. Visualizations were produced via Matplotlib version 3.10 and Seaborn version 0.13.

Per the AHA Journals Guidelines on AI usage, automated assistive writing technologies (ChatGPT and Claude) were used strictly as technical aids for Python code refactoring, reference formatting, and initial grammatical editing. The author assumes full responsibility for the content, scientific accuracy, and integrity of all analyses and written text presented in this study.

### 2.5 Independent Data Access and Analysis

As required by the American Heart Association Journals Research Guidelines, the corresponding author (Khalid Mohammed Al-Dhayani) attests to having full access to all the data in the study and takes sole responsibility for the integrity of the data, the accuracy of the data analysis, and the decision to submit the manuscript.

### 2.6 Human Subjects and Ethical Approval

This study analyzed publicly available, aggregated country-level secondary data obtained from the IHME GBD 2023 database and the World Bank Open Data repository. No individual patient identifiable information or direct human subject recruitment was involved. Institutional ethics approval and participant written informed consent were waived by the Institutional Review Board of the University of Amran in accordance with institutional guidelines for secondary public health data research.

### 2.7 Sex- and Demographic-Specific Data Statement

The American Heart Association Journals Research Guidelines on demographic reporting, primary exposures and outcomes in this ecological study were evaluated as national age-standardized population-level rates across 204 countries and territories. Sex-stratified and race/ethnicity-stratified analyses were not performed because primary exposure variables were analyzed as unified national risk-attributable mortality metrics. Overall, global socioeconomic disparities were evaluated via World Bank national income group classifications.

### 2.8 Data availability and reproducibility (TOP guidelines)

This study adheres to the transparency and openness promotion (TOP) guidelines. The underlying raw data are publicly available through the IHME GBD Results Tool (https://ghdx.healthdata.org/gbd-results-tool) and the World Bank Open Data repository (https://data.worldbank.org). The harmonized country-year analytical dataset generated during this study has been deposited in the Zenodo open repository and is publicly available at https://doi.org/10.5281/zenodo.21740644.

## 3. RESULTS

### 3.1 Study population and analytical dataset

The final analytical dataset included 204 countries and territories observed annually from 2000--2023, resulting in a balanced longitudinal panel comprising 4,896 country–year observations (204 countries and territories × 24 calendar years). All observations included in the primary analysis contained complete information on the outcome variable, cardiometabolic risk factors, and socioeconomic covariates. As the study was based entirely on aggregated national-level estimates, no individual participant recruitment, follow-up, or patient-level data were included.

### 3.2 Baseline characteristics and global socioeconomic differences

Substantial socioeconomic disparities in age-standardized CVD mortality were observed across the World Bank income groups (Table 1). High-income countries had the lowest mean CVD mortality rate (208.44 ± 101.89 deaths per 100,000 people), whereas progressively higher mortality rates were observed in upper-middle-income (302.69 ± 123.62), lower-middle-income (315.20 ± 160.67), and low-income countries (333.37 ± 148.13). A similar socioeconomic gradient was evident for mortality attributable to high SBP, with mean rates increasing from 115.39 ± 64.21 deaths per 100,000 people in high-income countries to 196.44 ± 92.35 in low-income countries. The GDP per capita also differed markedly between income groups, ranging from US$1,287.65 ± 753.09 in low-income countries to US$35,048.67 ± 27,470.97 in high-income countries (Table 1).

**Table 1.**
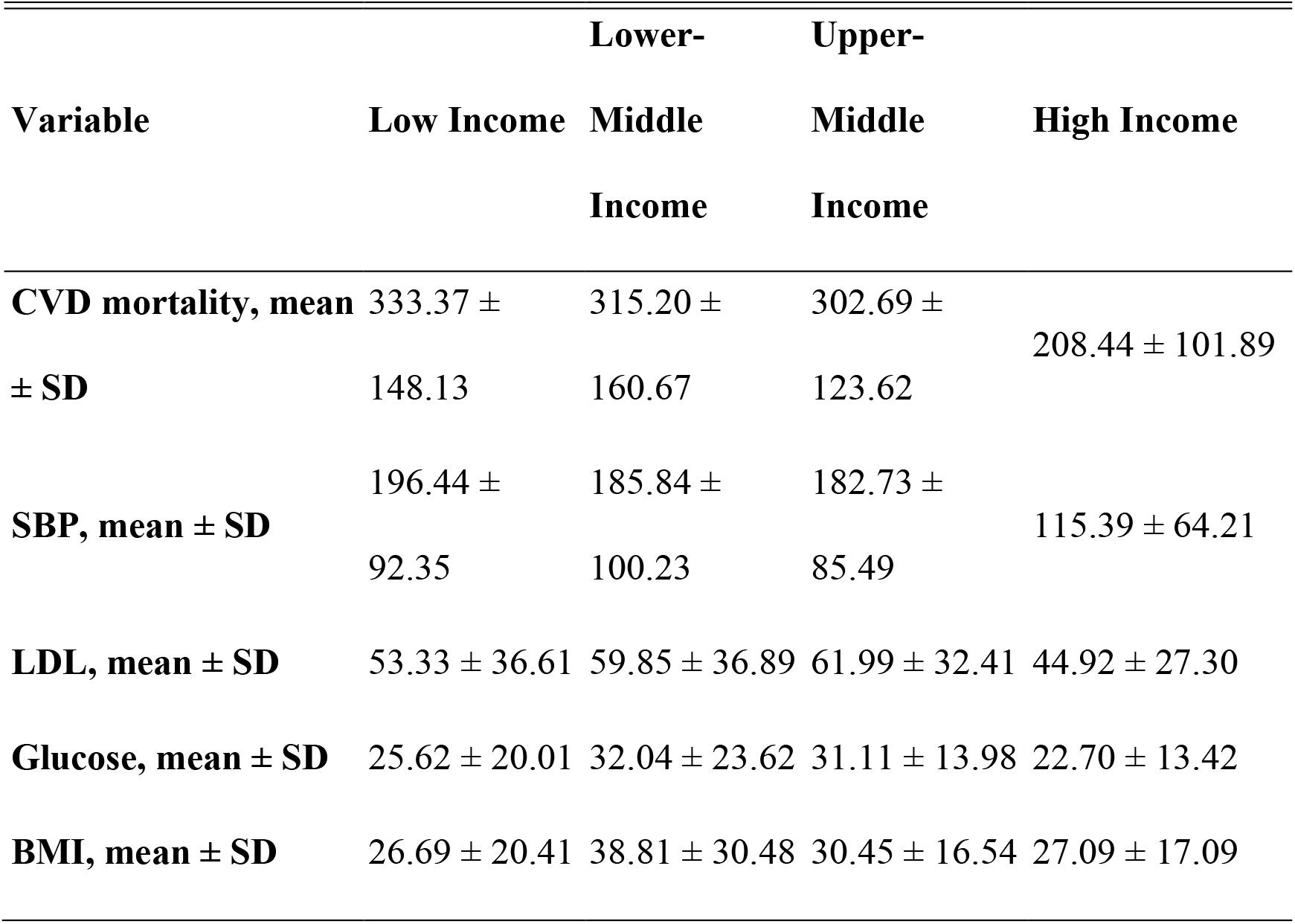
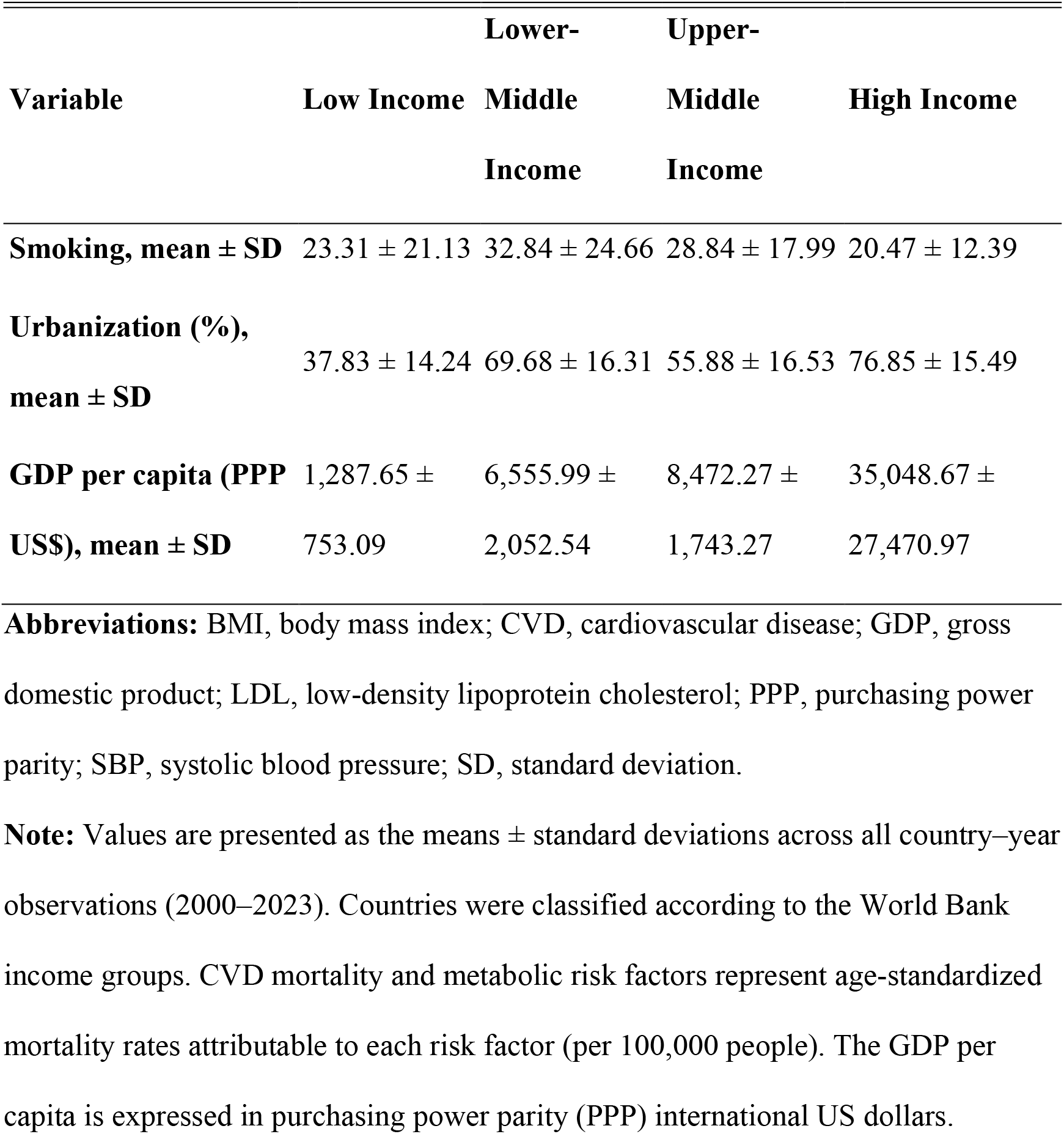
Summary Statistics of the Key Variables by World Bank Income Group (2000–2023)

Marked geographic variation was also observed. Small island developing states in Oceania recorded the highest average CVD mortality during the study period, including Nauru (892.48 deaths per 100,000 people), Tuvalu (725.54), and the Marshall Islands (708.39). Other countries with particularly high average mortality rates included Egypt (699.37), Sudan (683.32), Afghanistan (657.56), Morocco (620.18), Uzbekistan (612.83), Ukraine (592.24), and Turkmenistan (586.99) (Table S1). Most of these high-burden countries demonstrated statistically significant declines in mortality over time, whereas trends in the Marshall Islands and Egypt were not statistically significant.

The global geographic distribution demonstrated pronounced spatial heterogeneity, with the highest average mortality concentrated across Eastern Europe, Central Asia, North Africa, and Oceania, whereas substantially lower mortality rates were observed throughout Western Europe, North America, and East Asia (Figure 3). Comparisons between countries at the highest and lowest ends of the mortality distribution further highlighted the considerable international disparities in CVD burden (Figure 5).

### 3.3 Temporal Trends in CVD Mortality and Risk-Attributable Mortality

Global age-standardized CVD mortality declined substantially between 2000 and 2023, decreasing from approximately 340 deaths per 100,000 people in 2000 to fewer than 260 deaths per 100,000 people in 2023 (Figure 1). This long-term decline was briefly interrupted by a transient increase from 2020–2021, corresponding to the acute phase of the COVID-19 pandemic, before it returned to its downward trajectory from 2022–2023.

**Figure 1.**
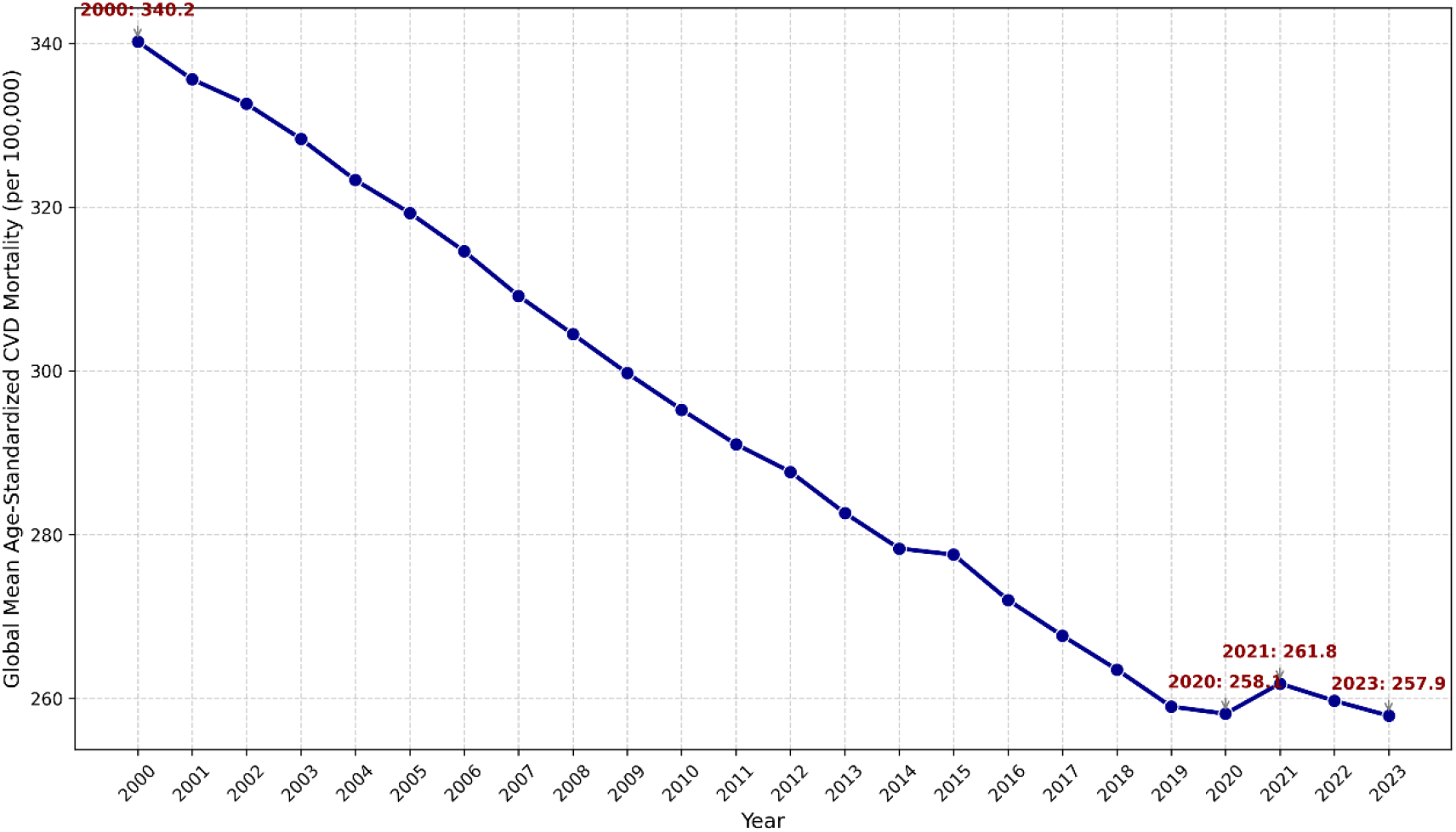
Global trends in the age-standardized cardiovascular disease (CVD) mortality rate (2000–2023). Line graph demonstrating the global mean age-standardized CVD mortality rate (deaths per 100,000 people) across 204 countries and territories from 2000--2023. The trend line reveals a steady secular decline, decreasing from approximately 340 deaths per 100,000 people in 2000 to fewer than 260 deaths per 100,000 people in 2023. A transient increase is observed from 2020–2021, corresponding to the acute phase of the COVID-19 pandemic, followed by a resumption of the downward trajectory in 2022 and 2023.

Mortality attributable to most major cardiometabolic and behavioral risk factors also declined over the study period (Figure 2). Smoking demonstrated the largest reduction, decreasing by 37.35%, from 34.66 to 21.71 deaths per 100,000 people. Mortality attributable to high LDL cholesterol declined by 30.00% (66.62 to 46.63), high SBP declined by 22.67% (196.35 to 151.83), and high fasting plasma glucose declined by 14.12% (30.78 to 26.43) (Table S3). In contrast, mortality attributable to high BMI increased slightly, by 1.38%, increasing from 32.78 to 33.23 deaths per 100,000 people.

**Figure 2.**
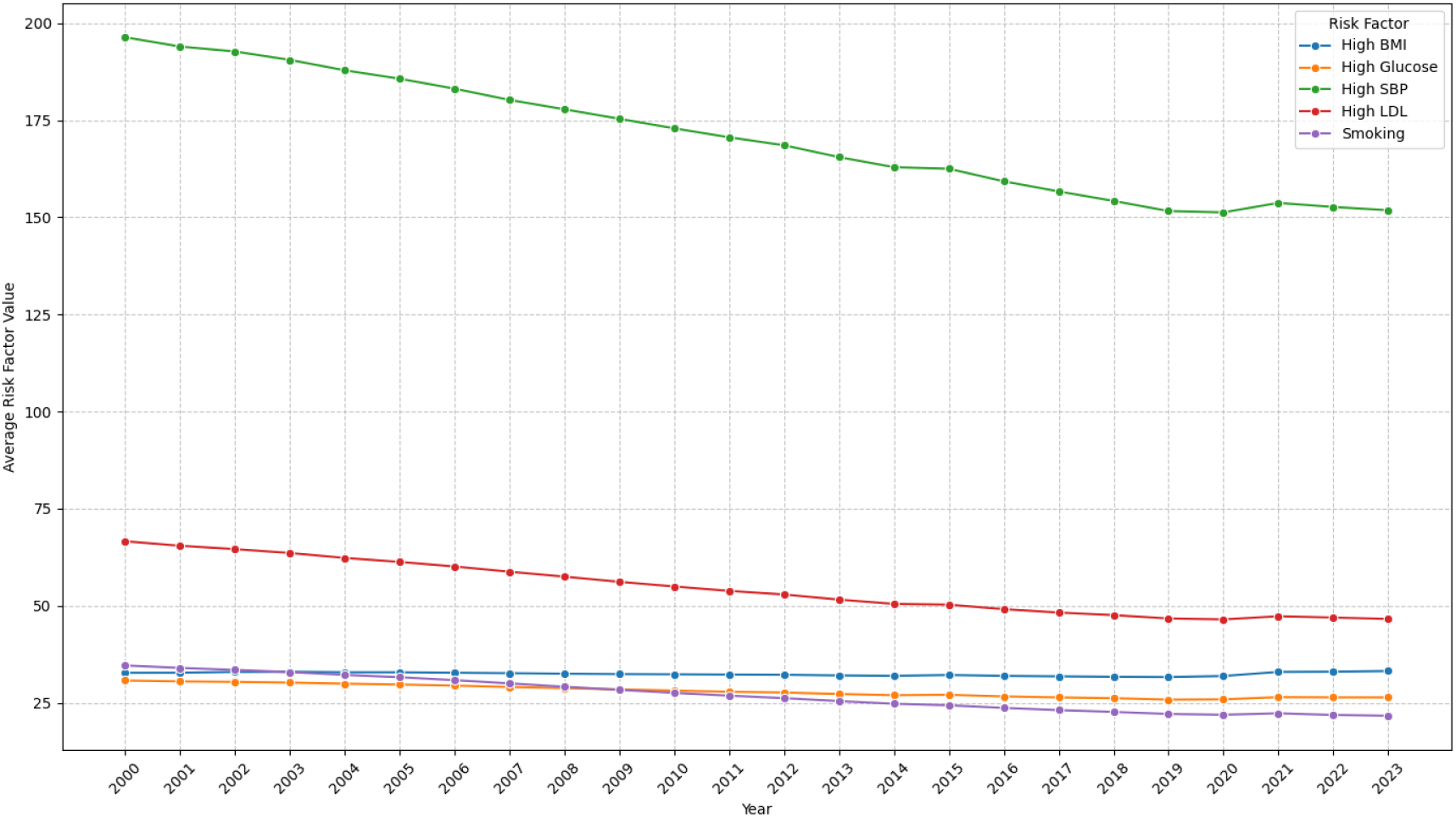
Global trends in age-standardized cardiovascular disease mortality attributable to major metabolic and behavioral risk factors (2000–2023). Multiline plot illustrating global mean age-standardized CVD mortality rates (deaths per 100,000 people) attributable to high systolic blood pressure (SBP), high low-density lipoprotein (LDL) cholesterol, high body mass index (BMI), high fasting plasma glucose, and smoking from 2000–2023. High SBP remained the leading risk factor throughout the study period. Decreasing trends are observed for SBP, LDL cholesterol, smoking, and fasting plasma glucose, whereas BMI-attributable mortality remains relatively stable, with a slight upward trajectory toward 2023. A synchronized increase across all risk factor pathways was evident during 2021, coinciding with the COVID-19 pandemic.

**Figure 3.**
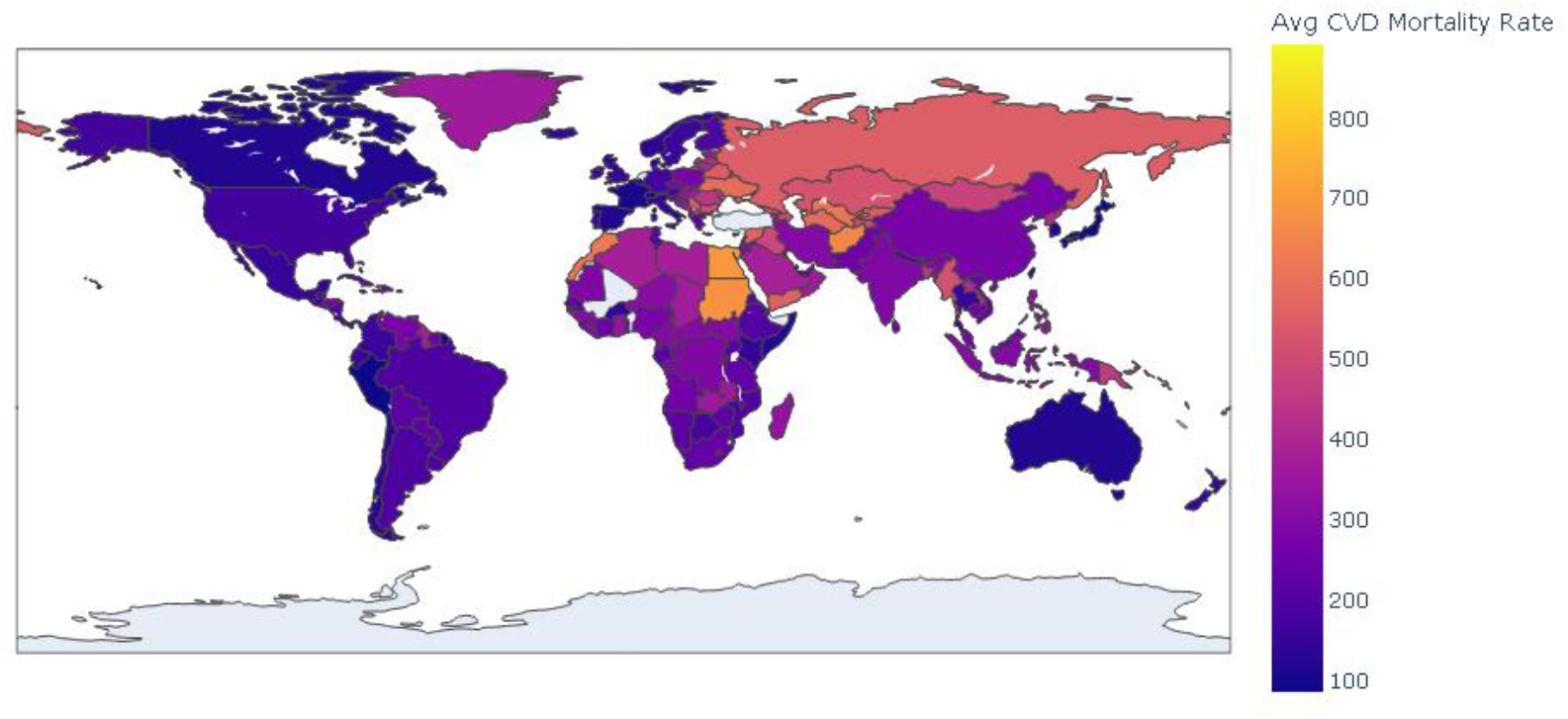
Global geographical distribution of average age-standardized cardiovascular disease mortality rates (2000–2023). Choropleth map illustrating the mean age-standardized CVD mortality rate across 204 countries and territories over the 24-year study period. Darker shades indicate lower average mortality (<200 deaths per 100,000 people), whereas lighter or warmer colors represent countries with the highest mortality burden (>600 deaths per 100,000 people), which are primarily concentrated in Eastern Europe, Central Asia, North Africa, and Oceania.

Annual percentage change analyses revealed a temporary reversal of declining trends from 2020–2021 across several risk factor pathways, with increases observed particularly in 2021, followed by renewed declines from 2022–2023 (Table S2). Similar temporal patterns were observed across representative countries, including a synchronized increase during 2021 followed by subsequent reductions (Figure S4). The geographic distribution of SBP-attributable mortality closely mirrored the distribution of overall CVD mortality (Figure 4).

**Figure 4.**
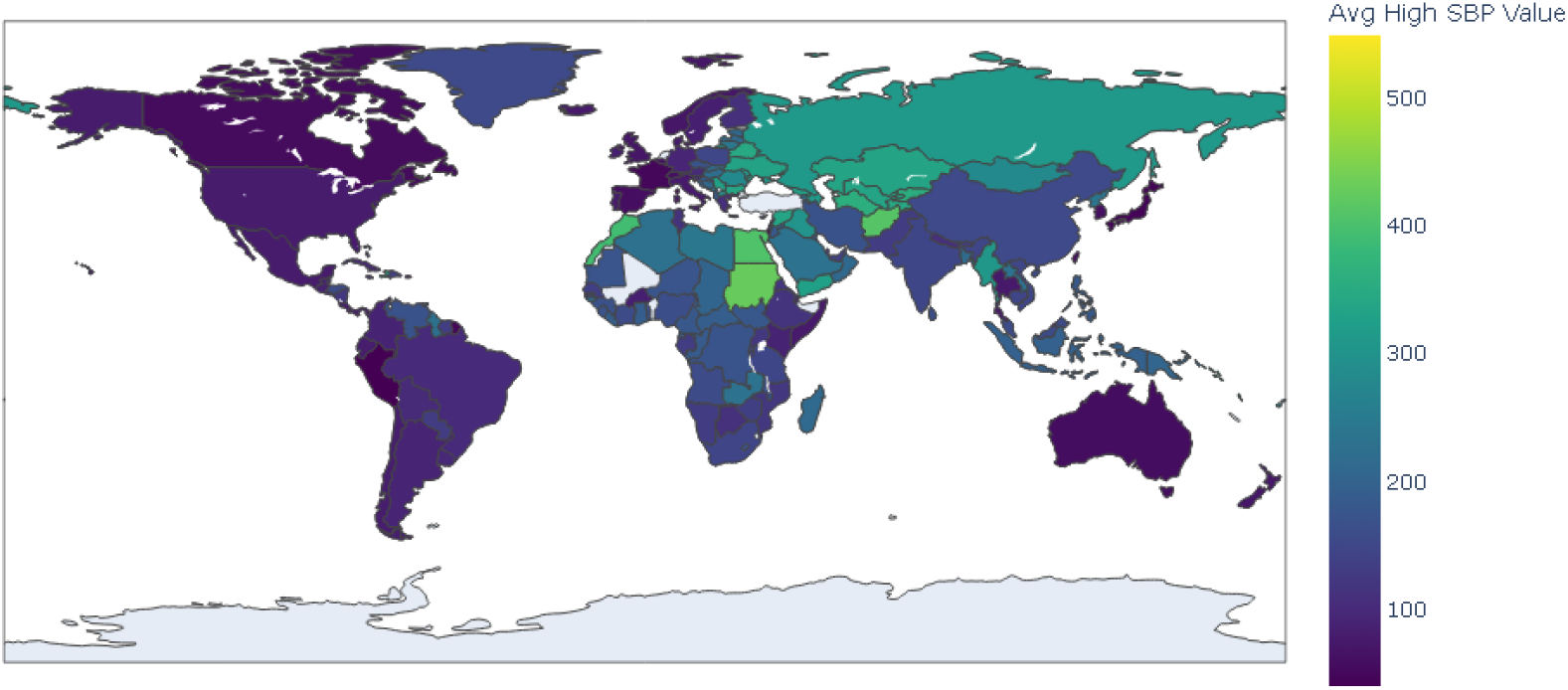
Global geographical distribution of average cardiovascular disease mortality attributable to high systolic blood pressure (2000–2023). Choropleth map illustrating the mean age-standardized CVD mortality rate attributable to high systolic blood pressure across 204 countries and territories from 2000–2023. The spatial distribution of SBP-attributable mortality closely parallels the global pattern observed for overall CVD mortality.

**Figure 5.**
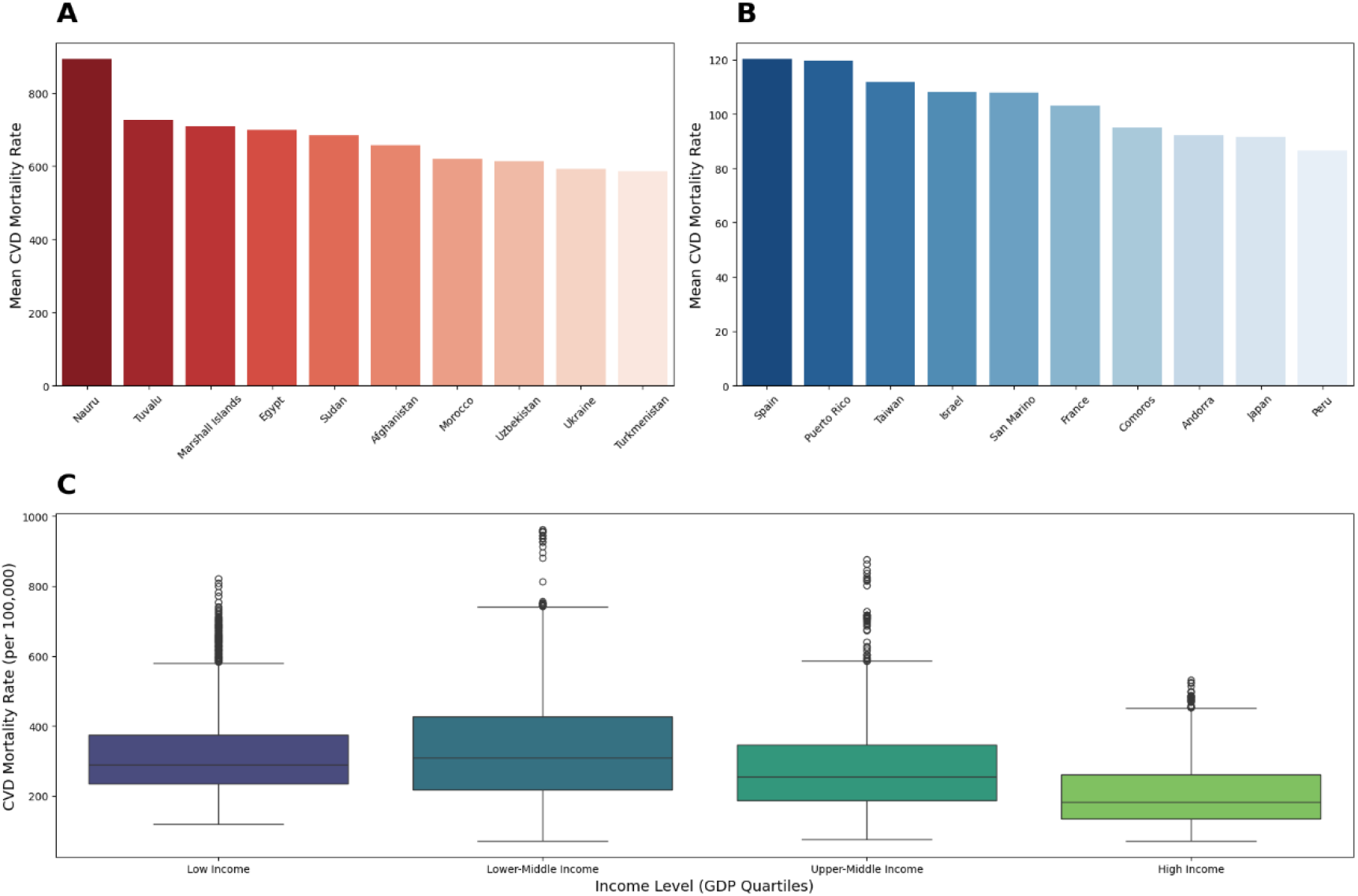
Geographic and socioeconomic disparities in global age-standardized cardiovascular disease mortality (2000–2023). Composite figure illustrating geographic and socioeconomic disparities in country-level CVD mortality burden. **(A)** The top ten countries with the highest mean age-standardized CVD mortality rates, led by Nauru, Tuvalu, and the Marshall Islands. **(B)** The top ten countries with the lowest mean age-standardized CVD mortality rates, led by Spain, Puerto Rico, and Taiwan. **(C)** Boxplots showing the distribution of CVD mortality according to World Bank income classifications, demonstrating a pronounced socioeconomic gradient in mortality burden.

### 3.4 Pairwise Correlations and Univariable Associations

Unadjusted correlation analyses revealed strong positive associations between age-standardized CVD mortality and major cardiometabolic risk factors (Figure 6). The strongest correlation was observed for high SBP (Pearson r = 0.99, P < 0.001), followed by high LDL cholesterol (r = 0.92, P < 0.001), high fasting plasma glucose (r = 0.82, P < 0.001), and high BMI (r = 0.79, P < 0.001). The GDP per capita demonstrated a moderate inverse correlation with CVD mortality (r = −0.33, P < 0.001).

**Figure 6.**
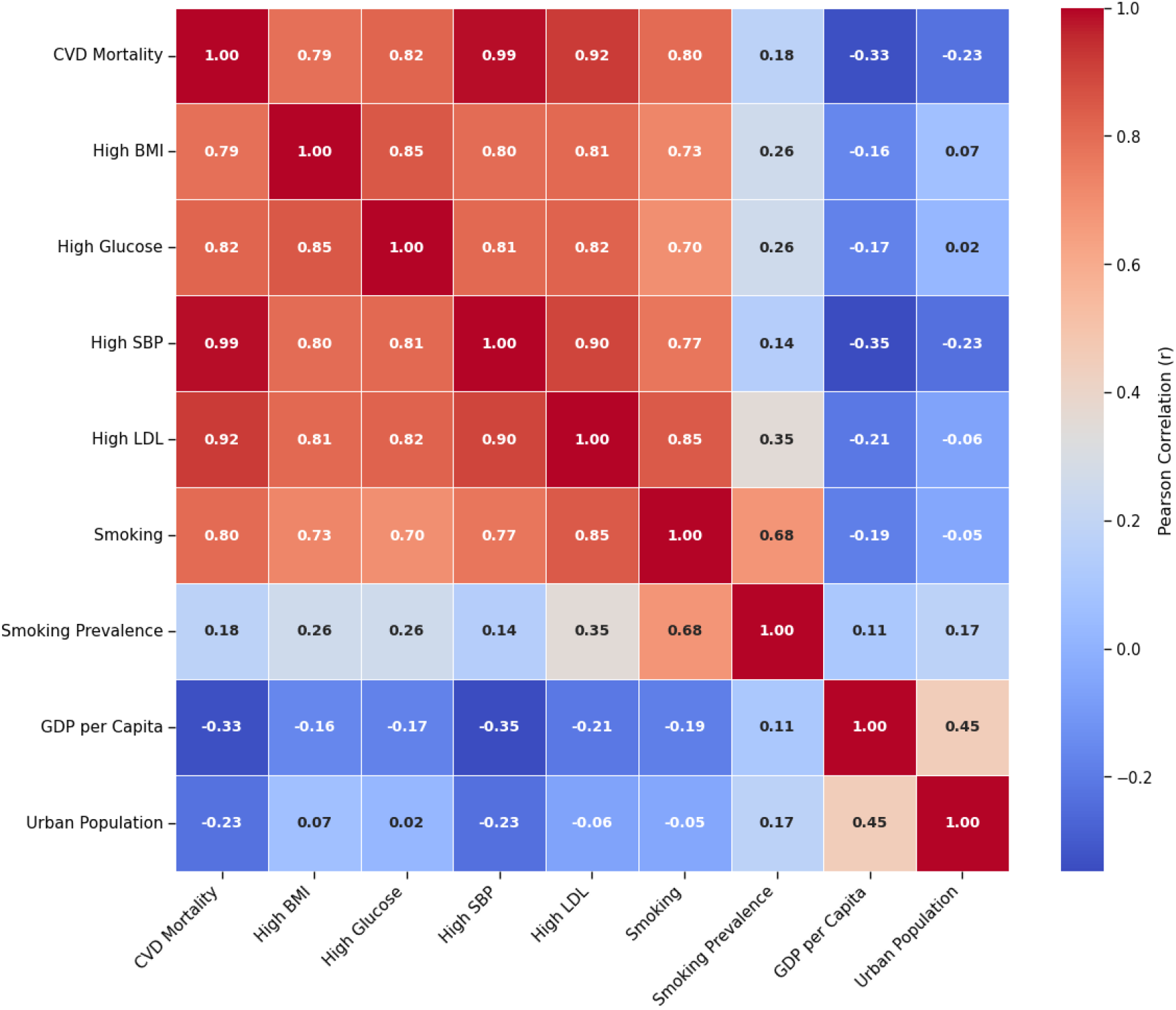
Pearson correlation heatmap of cardiovascular disease mortality, metabolic risk factors, and socioeconomic indicators. Heatmap displaying pairwise Pearson correlation coefficients (r) among age-standardized CVD mortality, cardiometabolic risk factors, and socioeconomic indicators. CVD mortality demonstrated an almost perfect positive correlation with high systolic blood pressure (r = 0.99), together with strong positive correlations with high LDL cholesterol (r = 0.92), high fasting plasma glucose (r = 0.82), and high BMI (r = 0.79).

Univariable regression analyses confirmed strong positive linear relationships between CVD mortality and each cardiometabolic risk factor, with high SBP exhibiting the strongest association (Figure 7). These findings also demonstrated substantial collinearity among cardiometabolic exposures, supporting the use of multivariable panel regression models.

**Figure 7.**
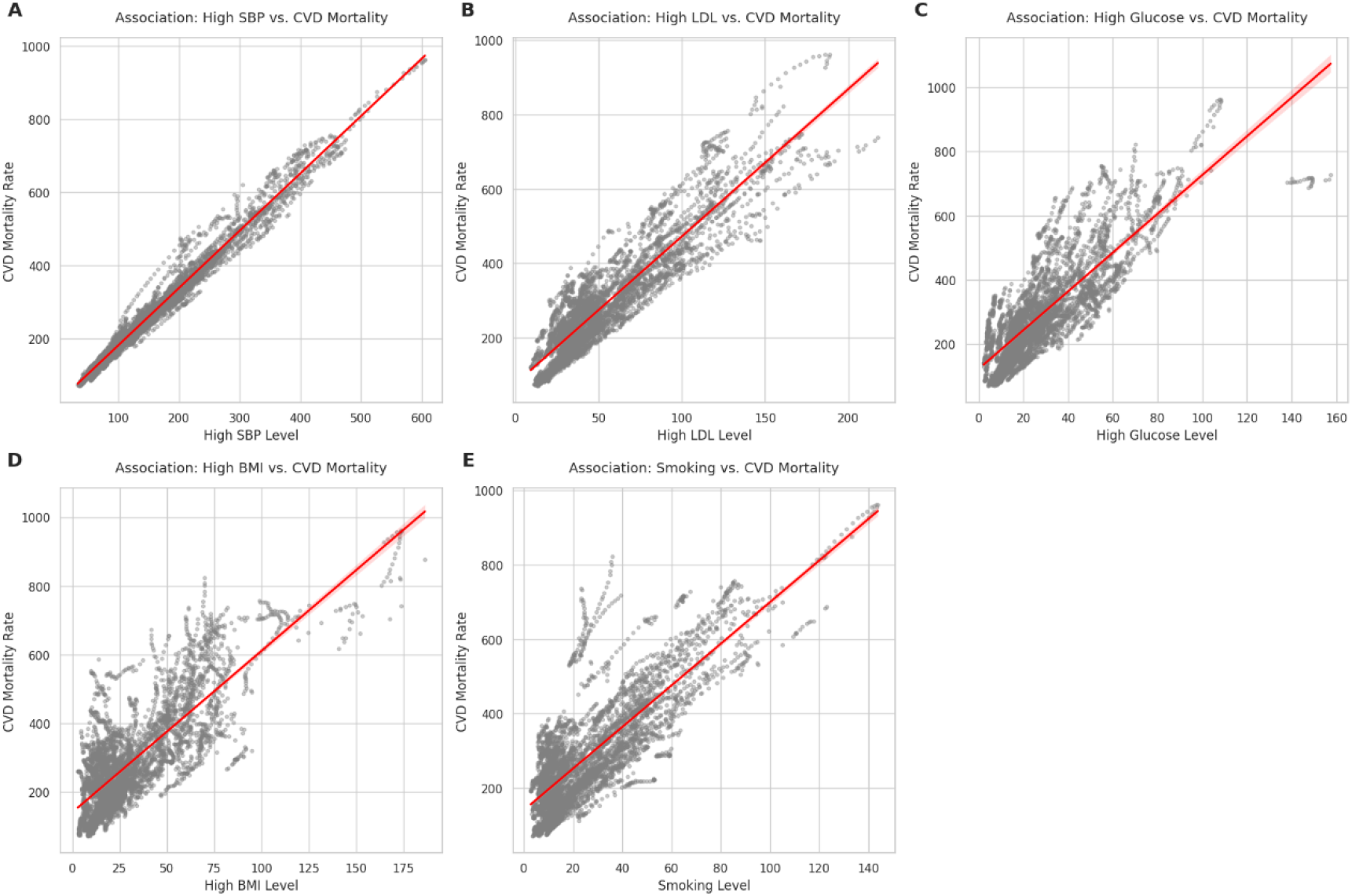
Univariable associations between individual metabolic and behavioral risk factors and cardiovascular disease mortality. Scatter plots with fitted linear regression lines and 95% confidence intervals illustrating the univariable associations between age-standardized CVD mortality and each major risk factor across 4,896 country-year observations: **(A)** high systolic blood pressure, **(B)** high LDL cholesterol, **(C)** high fasting plasma glucose, **(D)** high body mass index, and **(E)** smoking.

### 3.5 Primary Two-way Fixed-Effects Analysis

The primary multivariable analysis employed a TWFE panel regression model incorporating both country and calendar-year fixed effects with Driscoll–Kraay robust standard errors (Table 2).

**Table 2.** Primary two-way fixed-effects regression model examining associations between age-standardized cardiovascular disease mortality and major risk factors, 2000–2023.

| Variable | $\beta$ Coefficient | Standard Error | 95% Confidence Interval | *P* Value |
| --- | --- | --- | --- | --- |
| High SBP | 1.129 | 0.022 | 1.086 to 1.171 | <0.001 |
| High LDL | 0.622 | 0.020 | 0.583 to 0.662 | <0.001 |
| cholesterol |  |  |  |  |
| High fasting | 0.410 | 0.073 | 0.267 to 0.553 | <0.001 |
| plasma glucose |  |  |  |  |
| High BMI | 0.049 | 0.061 | −0.071 to 0.170 | 0.421 |
| Smoking | 0.452 | 0.026 | 0.401 to 0.502 | <0.001 |
| GDP per capita | −0.000053 | 0.000027 | −0.000106 to | 0.052 |
|  |  |  | 0.0000005 |  |
| Urban | −0.051 | 0.035 | −0.119 to 0.017 | 0.143 |
| population (%) |  |  |  |  |

Abbreviations: $\beta$ , regression coefficient; CI, confidence interval; LDL, low-density lipoprotein cholesterol; SBP, systolic blood pressure; BMI, high body mass; GDP, gross domestic product.
Notes: Values are regression coefficients from the primary two-way fixed-effects model adjusted for country and calendar year fixed effects. Age-standardized cardiovascular disease mortality (deaths per 100,000 people) was the dependent variable. Two-sided P values <0.05 were considered statistically significant.

High SBP had the strongest independent association with age-standardized CVD mortality (β = 1.129, 95% CI: 1.086–1.171, P < 0.001). Significant positive associations were also observed for high LDL cholesterol (β = 0.622, 95% CI: 0.583–0.662, P < 0.001), high fasting plasma glucose (β = 0.410, 95% CI: 0.267–0.553, P < 0.001), and smoking (β = 0.452, 95% CI: 0.401–0.502, P < 0.001).

After adjustment for other risk factors and fixed effects, high BMI was not significantly associated with CVD mortality (β = 0.049, 95% CI: −0.071 to 0.170, P = 0.421). The GDP per capita demonstrated a modest inverse association that approached statistical significance (β = −0.000053, 95% CI: −0.000106 to 0.0000005, P = 0.052), whereas urbanization was not significantly associated with mortality (β = −0.051, 95% CI: −0.119 to 0.017, P = 0.143).

Overall, after controlling for both country-specific characteristics and global temporal effects, elevated SBP remained the dominant population-level determinant of CVD mortality, followed by LDL cholesterol, smoking, and fasting plasma glucose (Table 2; Figure 8).

**Figure 8.**
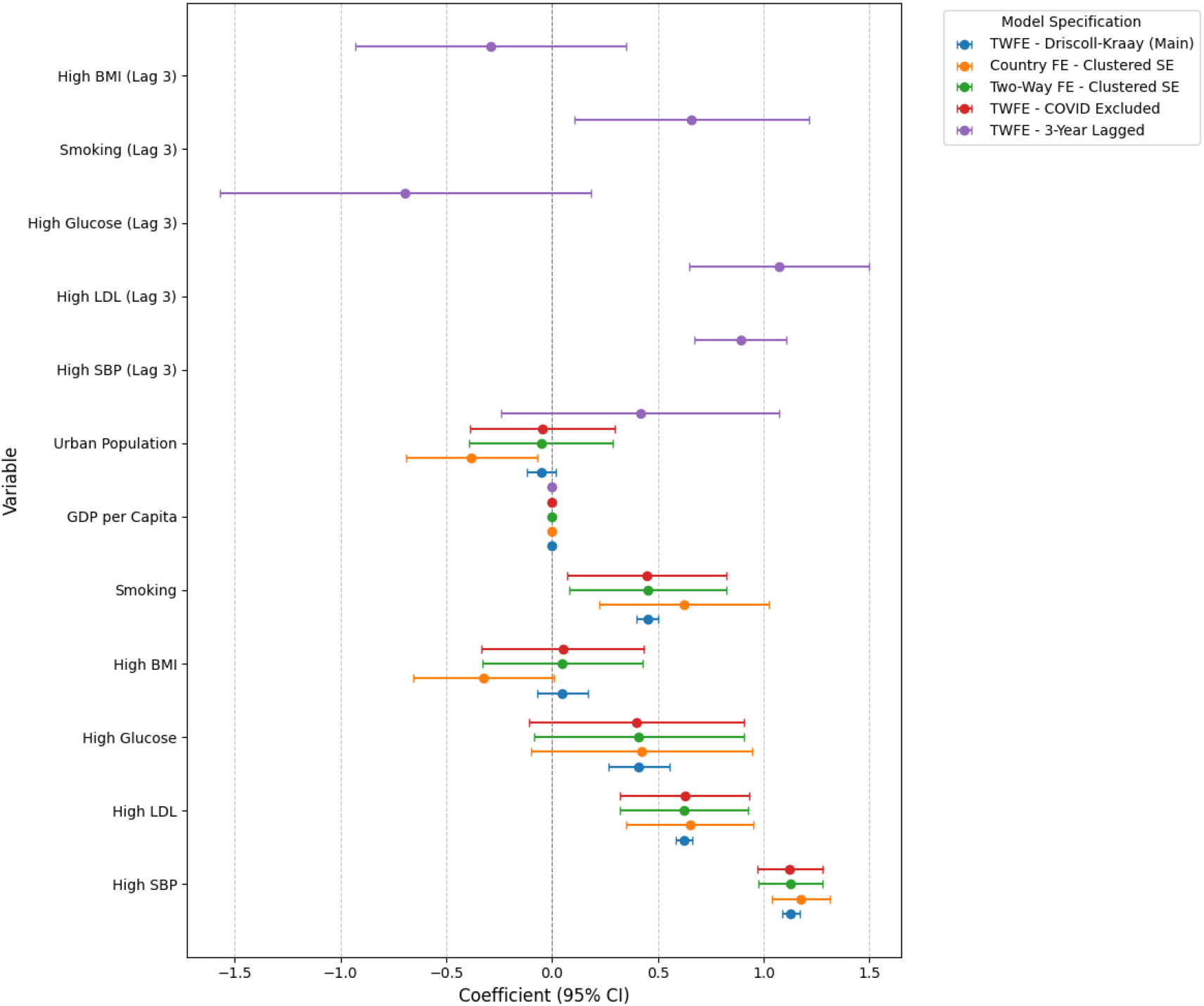
Forest plot of adjusted longitudinal population-level associations between cardiometabolic risk factors and cardiovascular disease mortality across multiple model specifications. Coefficient plot comparing regression estimates (β) and 95% confidence intervals from the primary two-way fixed-effects (TWFE) model with Driscoll–Kraay standard errors against multiple sensitivity analyses, including country fixed effects, TWFEs with clustered standard errors, exclusion of the COVID-19 pandemic years (2020–2021), and models incorporating 3-year lagged exposures. High systolic blood pressure, high LDL cholesterol, and smoking status demonstrated robust positive

### 3.6 Sensitivity Analysis Comparing Country Fixed Effects and Two-Way Fixed Effects

A comparison of the country fixed effects model with the TWFE specification demonstrated considerable stability in the estimated associations (Table 3). The regression coefficient for high SBP changed only slightly from 1.176--1.129, whereas the coefficients for high LDL cholesterol decreased from 0.652--0.622, and those for high fasting plasma glucose decreased from 0.425--0.410. Smoking remained positively associated with mortality, although its coefficient decreased from 0.623 to 0.452.

**Table 3.**
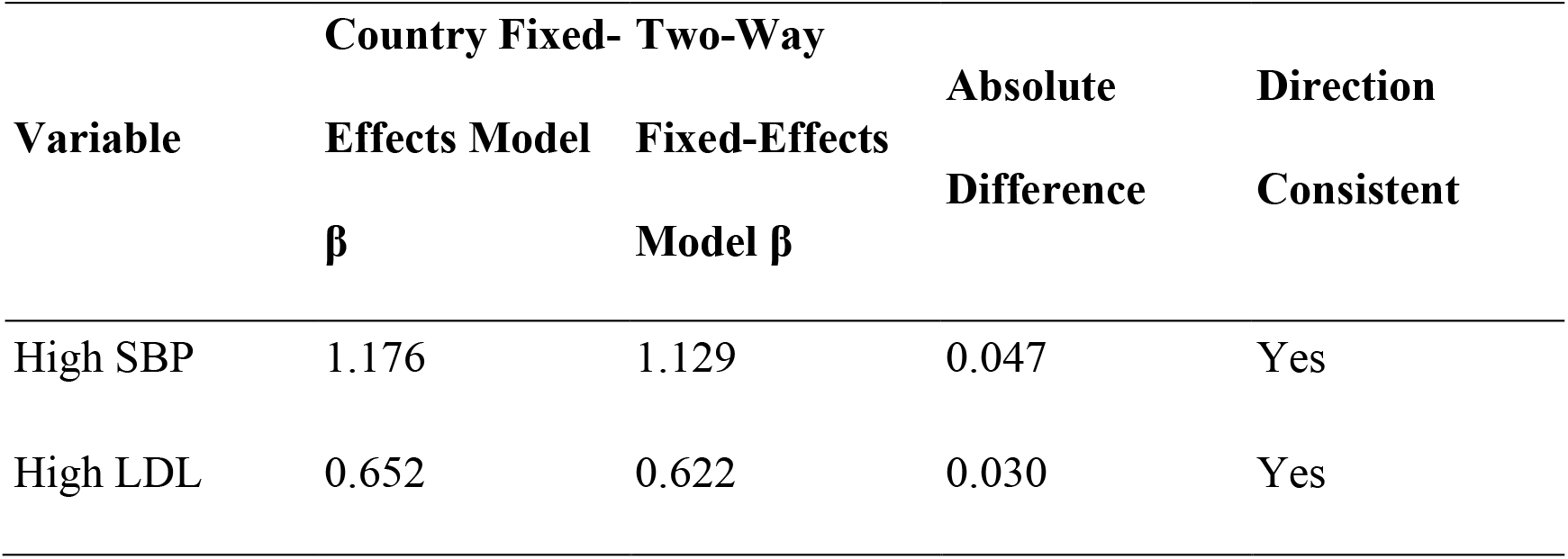
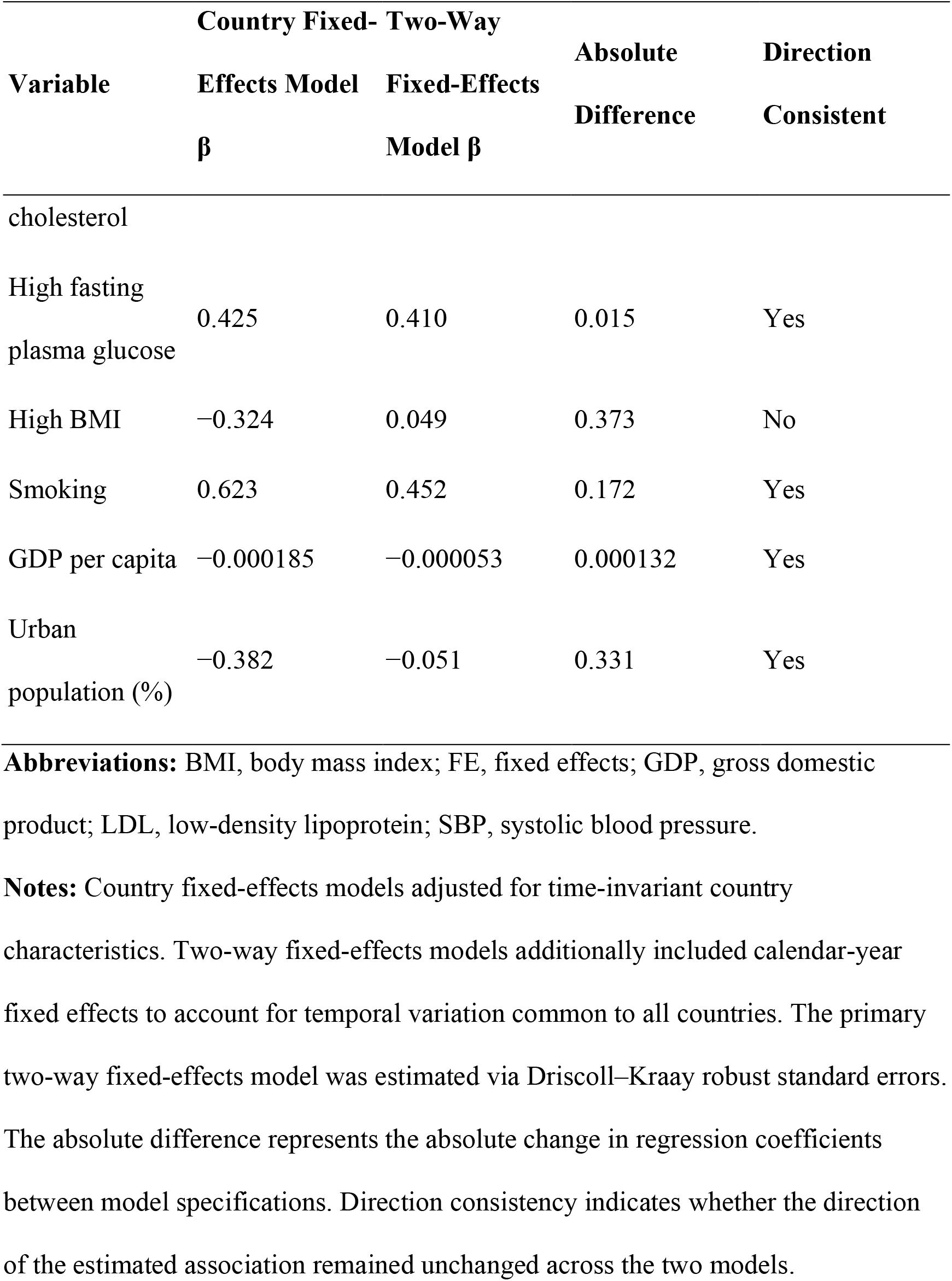
Sensitivity analysis comparing country fixed effects and two-way fixed effects regression models for associations between age-standardized cardiovascular disease mortality and major risk factors, 2000–2023.

The association for high BMI was particularly sensitive to adjustment for calendar-year effects. The coefficient changed from negative under the country fixed effects model (β = −0.324) to slightly positive under the TWFE model (β = 0.049), representing an absolute change of 0.373. This finding suggests that the apparent negative association observed in simpler models was largely attributable to unaccounted temporal trends. Socioeconomic coefficients were also attenuated after adjustment for year effects while retaining their inverse direction.

### 3.7 COVID-19 exclusion sensitivity analysis

Excluding observations from 2020 and 2021 produced estimates that were highly consistent with the primary analysis (Table 4). High SBP was strongly associated with CVD mortality (β = 1.123, 95% CI: 0.969–1.277, P < 0.001), as were high LDL cholesterol (β = 0.626, 95% CI: 0.320–0.933, P < 0.001) and smoking (β = 0.447, 95% CI: 0.072–0.822, P = 0.019). High fasting plasma glucose remained positively associated but was no longer statistically significant (β = 0.400, P = 0.125), whereas high BMI remained close to the null value (β = 0.051, P = 0.795). These findings indicate that primary associations were not materially influenced by the COVID-19 pandemic.

**Table 4.**
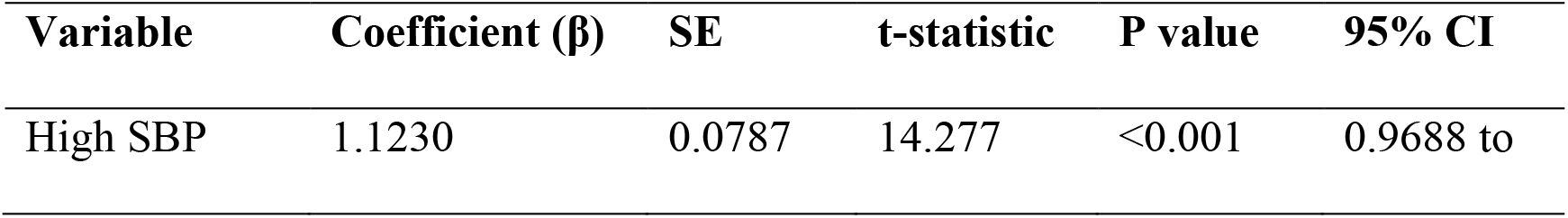
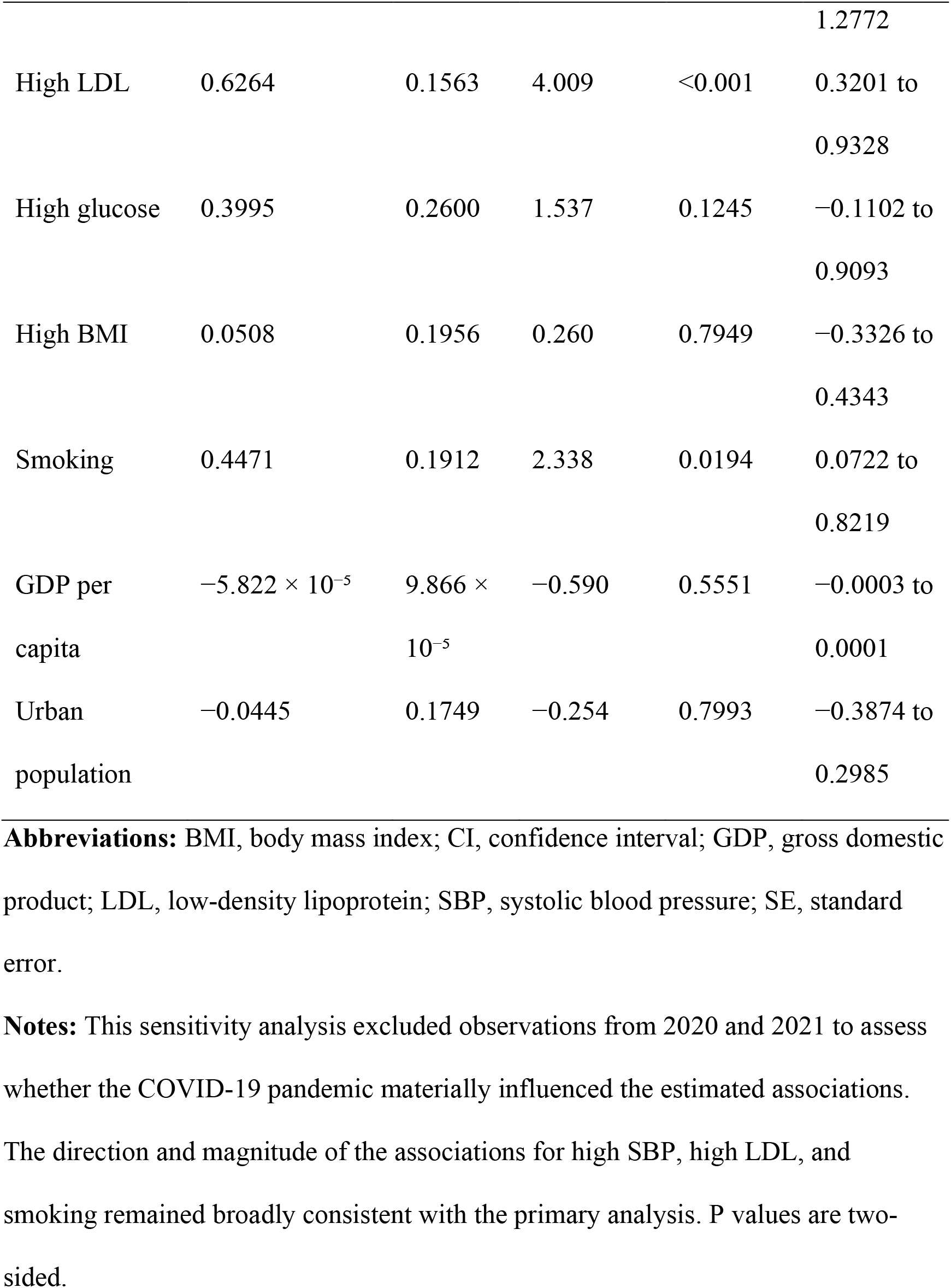
Sensitivity Analysis Excluding the COVID-19 Pandemic Years (2020–2021)

### 3.8 Three-Year Lagged Analysis

Analyses incorporating a three-year lag between exposure and outcome supported temporal precedence of the observed associations (Table 5). High SBP measured three years earlier remained strongly associated with subsequent CVD mortality (β = 0.891, 95% CI: 0.675–1.108, P < 0.001). Lagged high LDL cholesterol demonstrated an even stronger association (β = 1.073, 95% CI: 0.650–1.496, P < 0.001), whereas lagged smoking also remained significant (β = 0.659, 95% CI: 0.105–1.213, P = 0.020). The GDP per capita showed a significant inverse association with future mortality (β = −0.0003, P = 0.030), whereas urbanization remained nonsignificant.

**Table 5.**
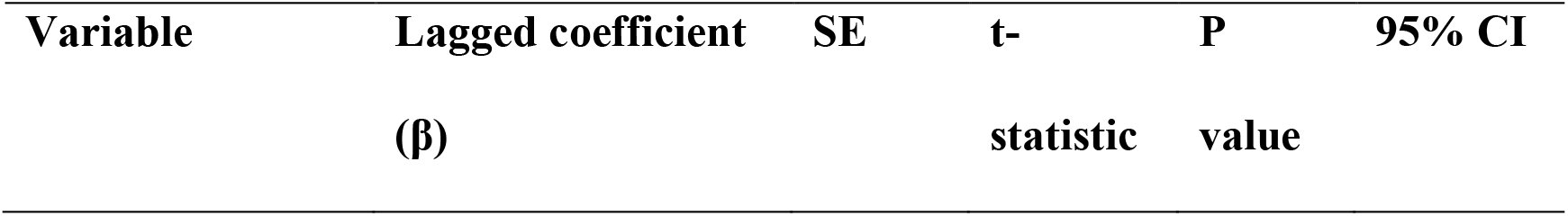
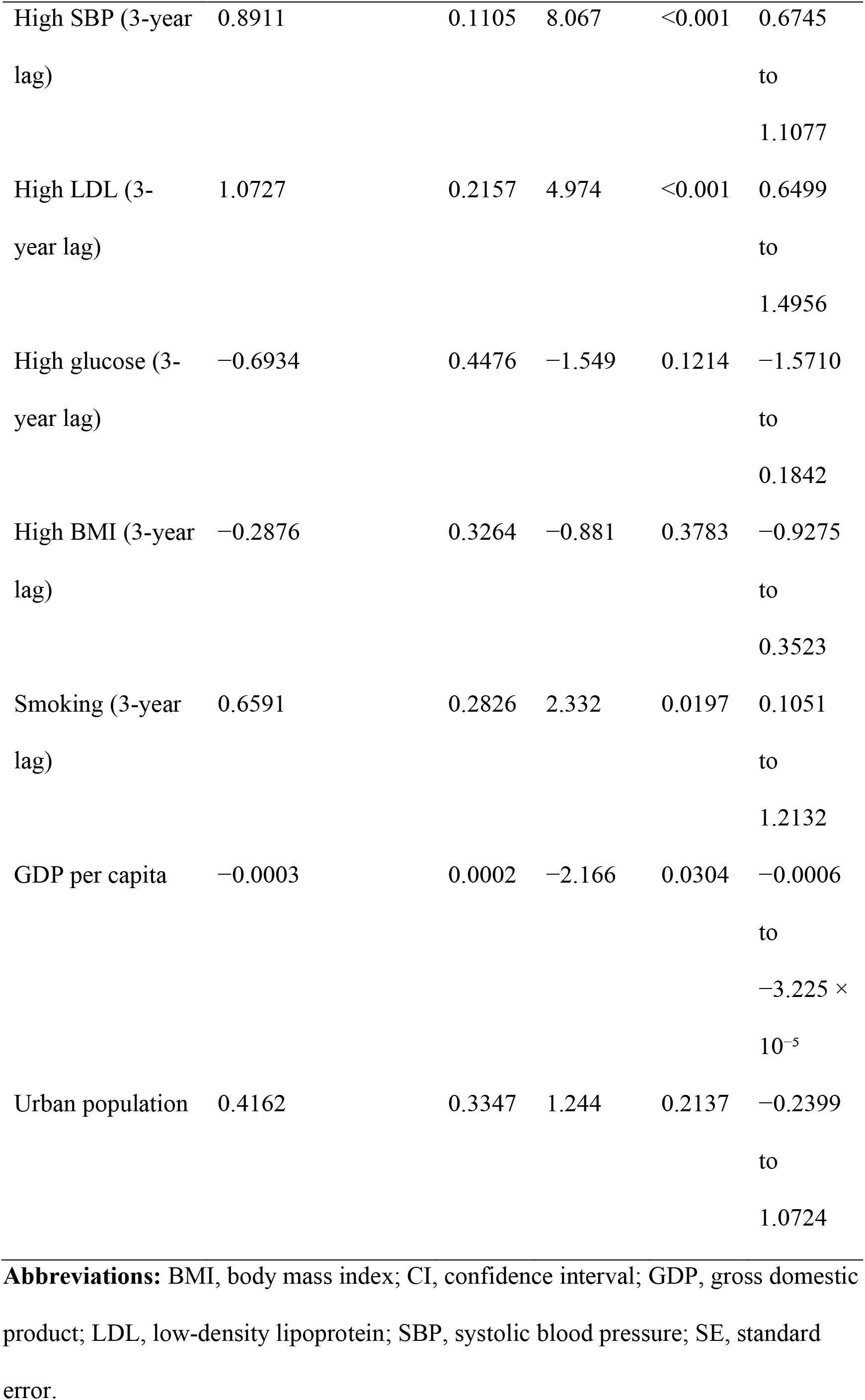
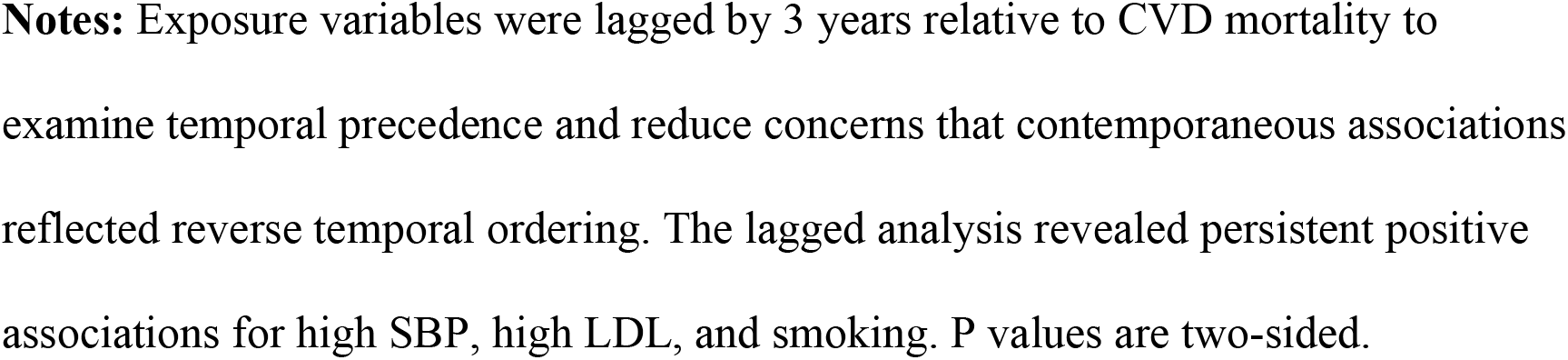
Three-Year Lagged Associations between Cardiovascular Disease Risk Factors and Mortality.

### 3.9 Assessment of Nonlinearity

Quadratic terms were incorporated to assess potential nonlinear exposure–response relationships (Table 6). Although all the linear associations remained positive, none of the quadratic terms reached statistical significance, including high SBP (P = 0.316), high LDL cholesterol (P = 0.569), high fasting plasma glucose (P = 0.567), high BMI (P = 0.122), and smoking (P = 0.581). These findings support the appropriateness of linear panel regression models across the observed exposure ranges.

**Table 6.**
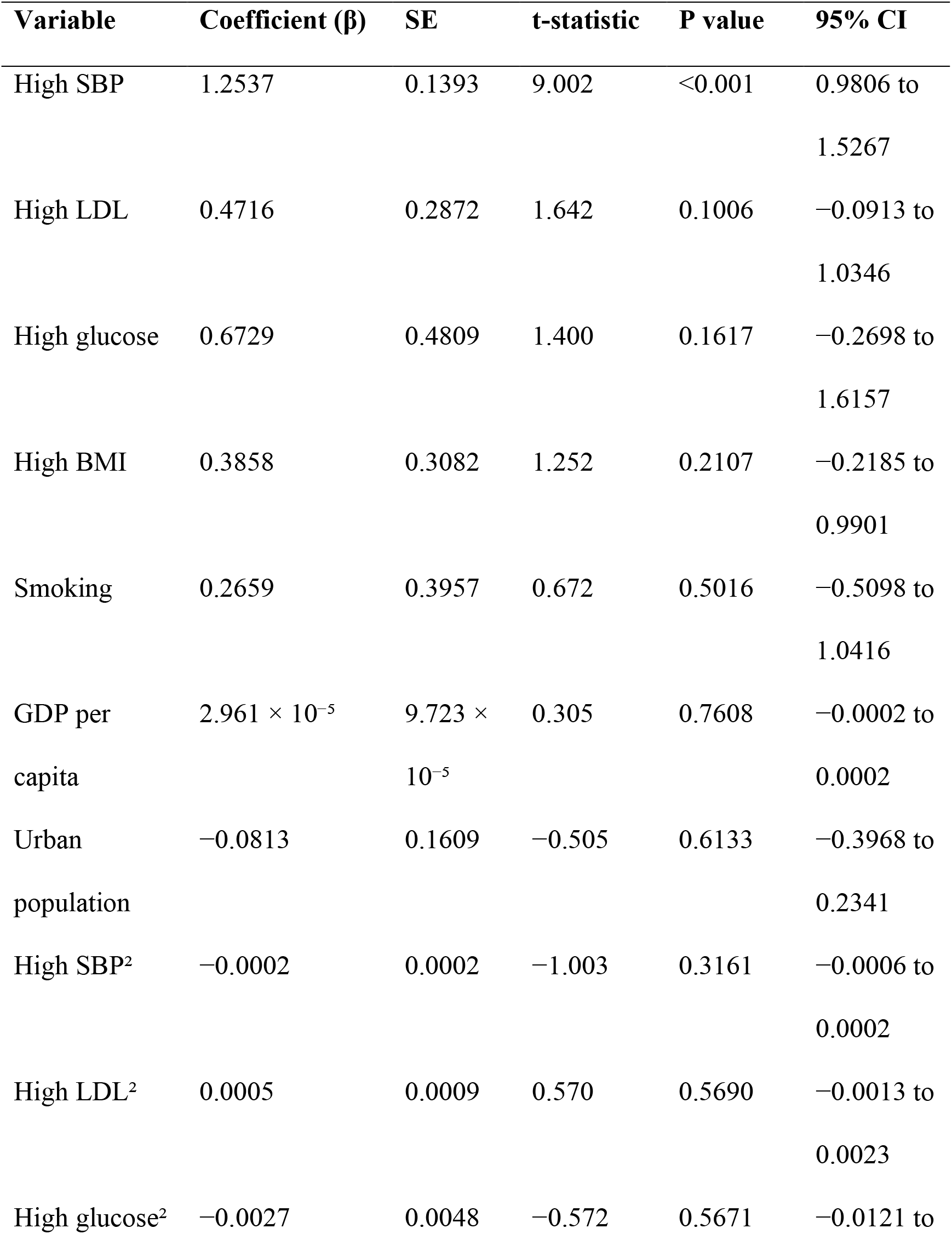
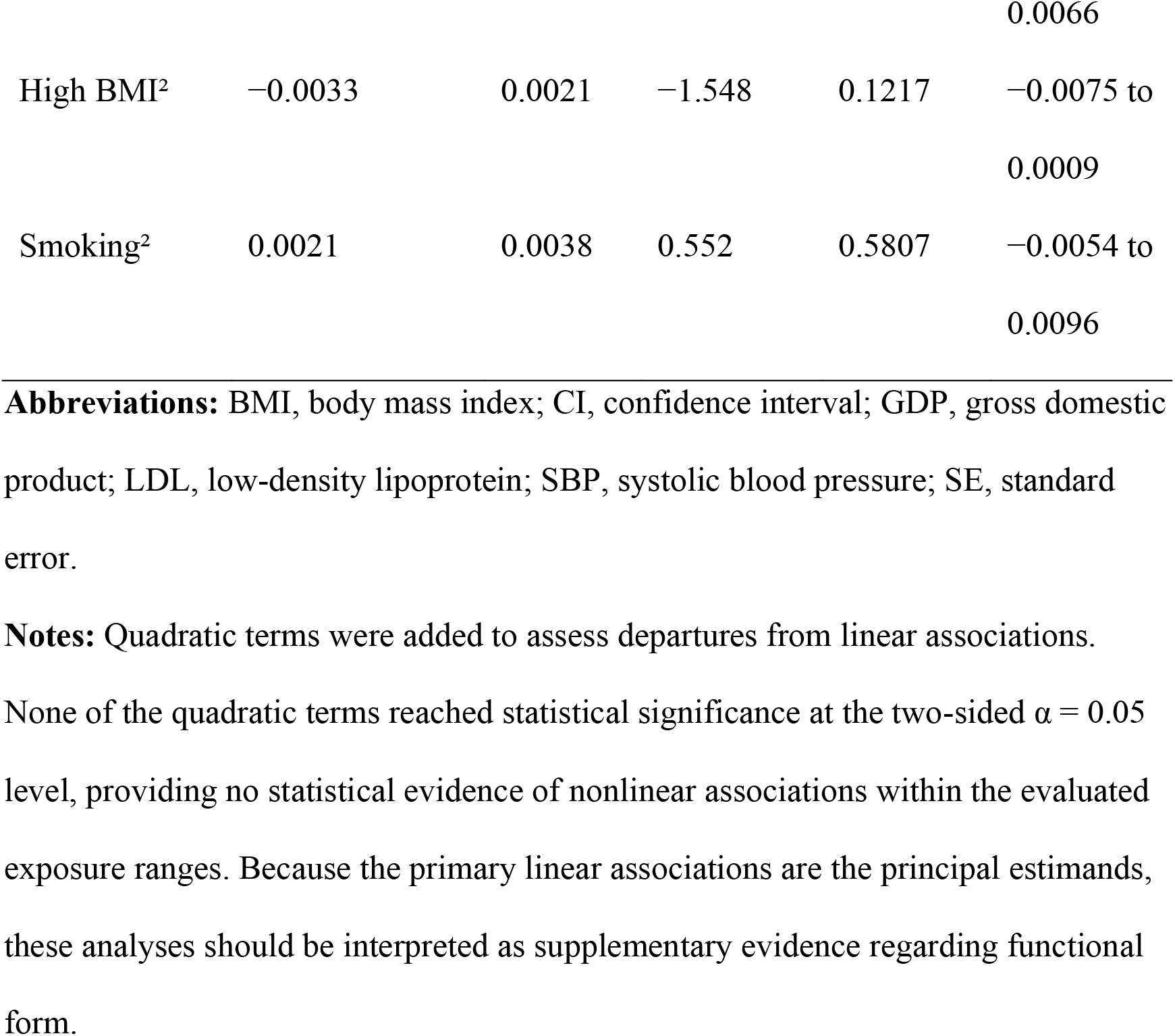
Assessment of Nonlinearity Using Quadratic Terms.

| Variable | Coefficient ($\beta$) | SE | t-statistic | P value | 95% CI |
| --- | --- | --- | --- | --- | --- |
| High SBP | 1.2537 | 0.1393 | 9.002 | <0.001 | 0.9806 to<br>1.5267 |
| High LDL | 0.4716 | 0.2872 | 1.642 | 0.1006 | −0.0913 to<br>1.0346 |
| High glucose | 0.6729 | 0.4809 | 1.400 | 0.1617 | −0.2698 to<br>1.6157 |
| High BMI | 0.3858 | 0.3082 | 1.252 | 0.2107 | −0.2185 to<br>0.9901 |
| Smoking | 0.2659 | 0.3957 | 0.672 | 0.5016 | −0.5098 to<br>1.0416 |
| GDP per<br>capita | $2.961 \times 10^{-5}$ | $9.723 \times 10^{-5}$ | 0.305 | 0.7608 | −0.0002 to<br>0.0002 |
| Urban<br>population | −0.0813 | 0.1609 | −0.505 | 0.6133 | −0.3968 to<br>0.2341 |
| High SBP<sup>2</sup> | −0.0002 | 0.0002 | −1.003 | 0.3161 | −0.0006 to<br>0.0002 |
| High LDL<sup>2</sup> | 0.0005 | 0.0009 | 0.570 | 0.5690 | −0.0013 to<br>0.0023 |
| High glucose<sup>2</sup> | −0.0027 | 0.0048 | −0.572 | 0.5671 | −0.0121 to |
|  |  |  |  |  | 0.0066 |
| High BMI<sup>2</sup> | −0.0033 | 0.0021 | −1.548 | 0.1217 | −0.0075 to<br>0.0009 |
| Smoking<sup>2</sup> | 0.0021 | 0.0038 | 0.552 | 0.5807 | −0.0054 to<br>0.0096 |

**Abbreviations:** BMI, body mass index; CI, confidence interval; GDP, gross domestic product; LDL, low-density lipoprotein; SBP, systolic blood pressure; SE, standard error.
**Notes:** Quadratic terms were added to assess departures from linear associations.
None of the quadratic terms reached statistical significance at the two-sided $\alpha = 0.05$ level, providing no statistical evidence of nonlinear associations within the evaluated exposure ranges. Because the primary linear associations are the principal estimands, these analyses should be interpreted as supplementary evidence regarding functional form.

### 3.10 Heterogeneity by country income group

Interaction analyses demonstrated significant heterogeneity across World Bank income groups (joint Wald test = 36.13, P = 0.00031) (Table 7). High SBP maintained consistently positive associations across all income categories (baseline β = 1.027, P < 0.001). High LDL cholesterol exhibited a significant negative interaction in upper-middle-income countries (β = −1.622, P = 0.0012), indicating a weaker association than in high-income countries. In contrast, smoking showed a significant positive interaction in upper-middle-income countries (β = 0.966, P = 0.039), suggesting a stronger population-level impact in these settings.

**Table 7.**
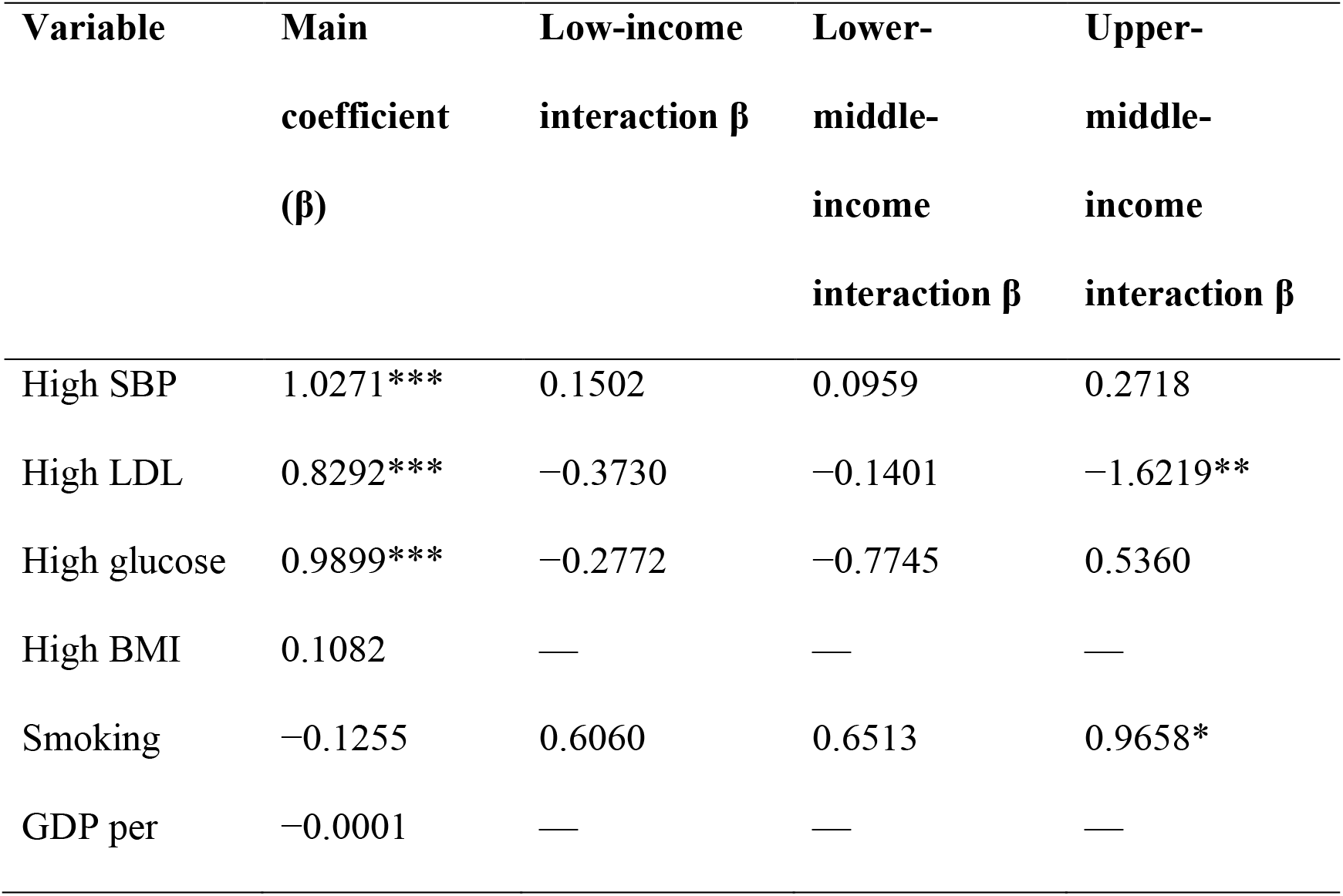
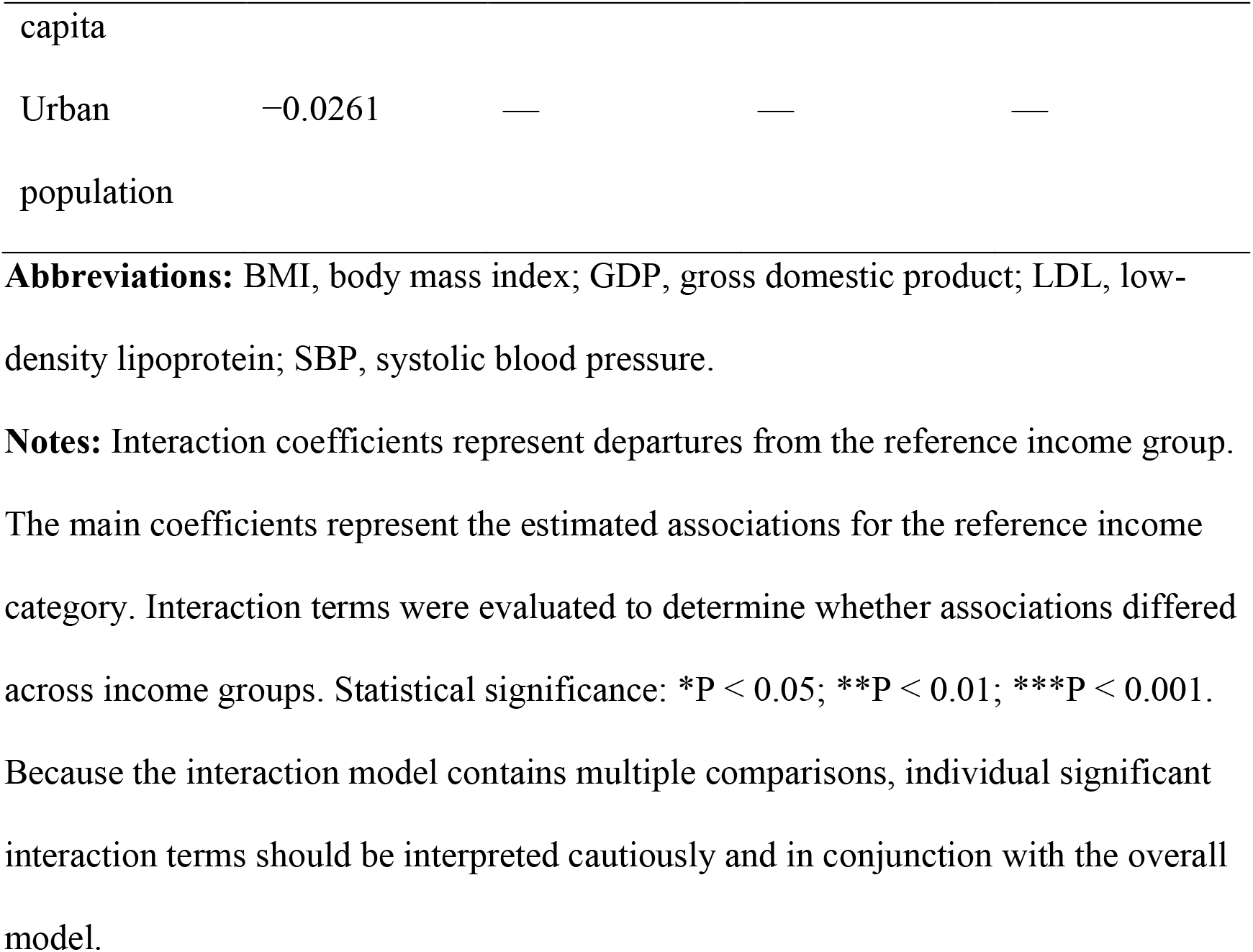
Heterogeneity of Associations by Country Income Group.

### 3.11 Heterogeneity by World Health Organization Region

Regional interaction models also demonstrated significant heterogeneity (joint Wald test = 47.02, P = 0.00058) (Table 8). High SBP remained positively associated with CVD mortality across all six WHO regions, with total coefficients ranging from 0.958 in the African Region (AFRO) to 1.308 in the Region of the Americas (AMRO). The most pronounced regional effect modification was observed for smoking, with significant positive interactions in AFRO (β = 0.833, P = 0.008), AMRO (β = 1.536, P = 0.002), and the western Pacific region (WPRO) (β = 1.299, P = 0.005). High LDL cholesterol demonstrated a particularly strong association with AFRO (total β = 1.458) compared with the European Region (EURO) baseline (β = 0.146).

**Table 8.** Heterogeneity of Associations by World Health Organization Region.

| Variable | Main<br>coefficient<br>($\beta$) | AFRO<br>interaction<br>n $\beta$ | AMRO<br>interaction<br>n $\beta$ | EMRO<br>interaction<br>n $\beta$ | SEARO<br>interaction<br>n $\beta$ | WPRO<br>interaction<br>n $\beta$ |
| --- | --- | --- | --- | --- | --- | --- |
| High<br>SBP | 1.2779**<br>* | −0.2447 | −0.1452 | −0.0244 | −0.1753 | 0.0365 |
| High<br>LDL | 0.4183* | 0.3194 | −0.1478 | 0.6319 | 0.5570 | −0.3002 |
| High<br>glucose | 0.7758* | −0.3698 | 0.0865 | −1.0482 | −0.9135 | −0.8797 |
| High<br>BMI | 0.0748 | — | — | — | — | — |
| Smoking | −0.3802 | 0.8332** | 1.5355** | 0.2786 | 0.6062 | 1.2989** |
| GDP per capita | −0.0001 | — | — | — | — | — |
| Urban population | −0.1263 | — | — | — | — | — |

**Abbreviations:** AFRO, African Region; AMRO, Region of the Americas; BMI, body mass index; EMRO, Eastern Mediterranean Region; GDP, gross domestic product; LDL, low-density lipoprotein; SBP, systolic blood pressure; SEARO, Southeast Asia Region; WPRO, Western Pacific Region.
**Notes:** Interaction coefficients represent differences in associations between the specified WHO region and the reference region. Statistical significance: \*P < 0.05; \*\*P < 0.01; \*\*\*P < 0.001. Significant positive interactions were observed for smoking in AFRO, AMRO, and WPRO. Interaction estimates should be interpreted as differences from the reference region rather than as region-specific total effects.

### 3.12 Influence and robustness analyses

Influence diagnostics identified 372 country-year observations (7.60% of the analytical sample) with Cook’s distance values exceeding the predefined threshold (4/n = 0.00082) (Table S4). These influential observations were concentrated primarily in small island developing states, including Micronesia and the Marshall Islands, as well as high-density countries such as Papua New Guinea and Bangladesh. Leave–one-country-out sensitivity analyses demonstrated excellent model stability, with exclusion of these countries producing only minimal changes in the principal TWFE coefficients (high SBP β range: 1.101–1.159; high LDL cholesterol β range: 0.621–0.671). These findings confirm that the primary results were not driven by individual high-leverage countries.

### 3.13 Model diagnostics and robust inference

Diagnostic testing demonstrated substantial violations of conventional regression assumptions. The Durbin–Watson statistic (0.0989) indicated strong positive autocorrelation, the Breusch–Pagan Lagrange multiplier test confirmed significant heteroskedasticity (statistic = 618.18, P < 0.001), and the Lilliefors test demonstrated nonnormal residuals (P = 0.001) (Table 9). These findings justify the use of Driscoll–Kraay robust standard errors, which provide valid statistical inference in the presence of autocorrelation, heteroskedasticity, and cross-sectional dependence.

**Table 9.**
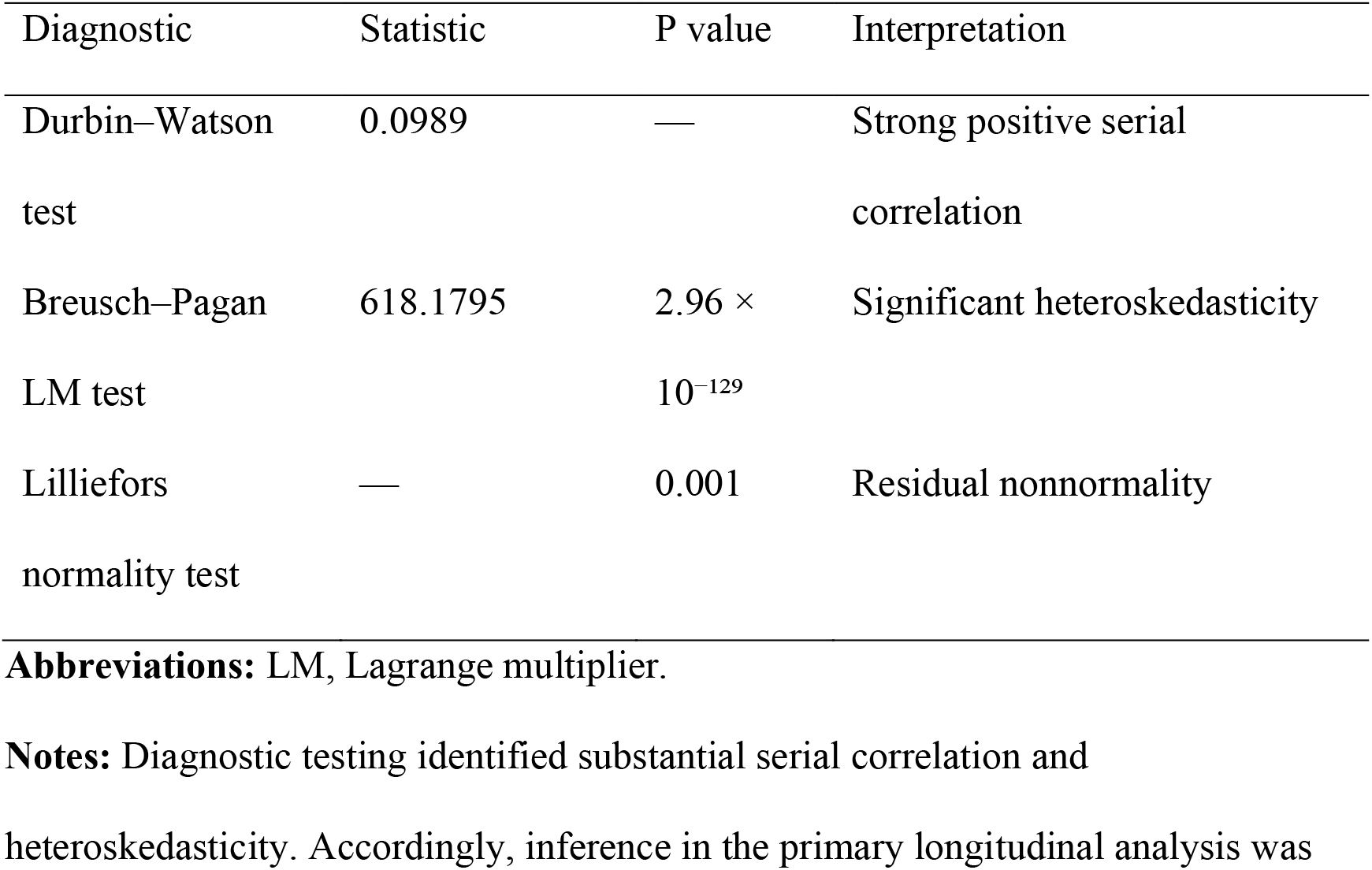
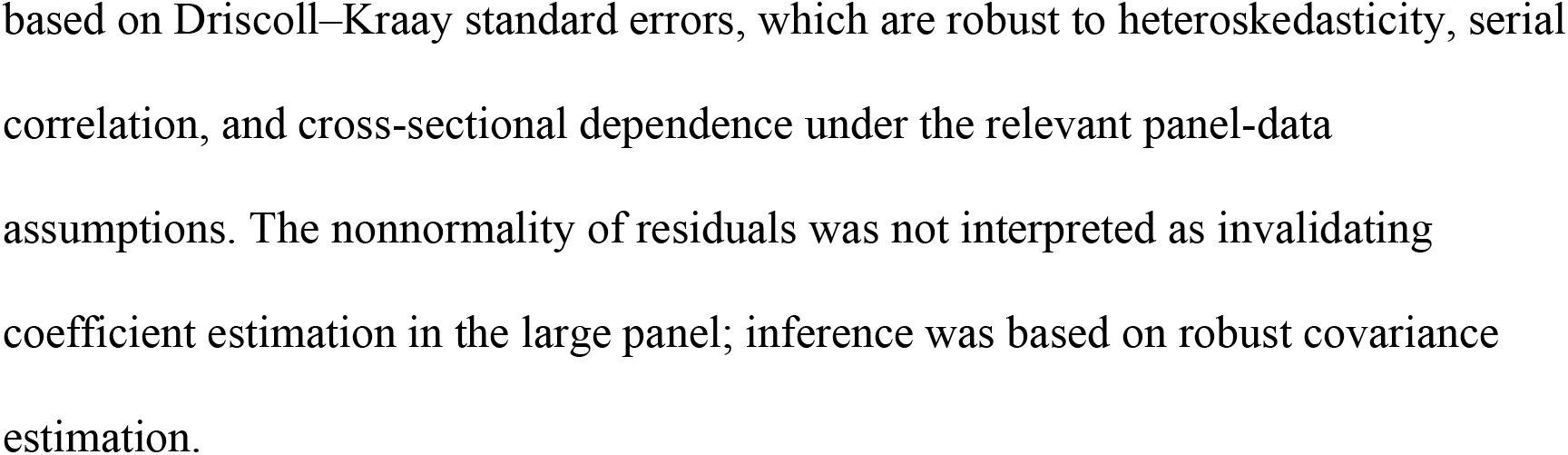
Model diagnostic tests.

### 3.14 Overall Pattern of Findings

Across the primary analyses, sensitivity analyses, and lagged models, elevated SBP consistently emerged as the strongest independent population-level predictor of age-standardized CVD mortality worldwide. High LDL cholesterol, high fasting plasma glucose, and smoking also maintained significant positive associations with mortality, whereas the association with high BMI was substantially attenuated after adjustment for other metabolic risk factors and fixed effects (Figure 8). Although global age-standardized CVD mortality declined considerably between 2000 and 2023, substantial international disparities persist, underscoring the continuing global importance of blood pressure control and comprehensive cardiometabolic risk reduction.

## 4. DISCUSSION

### 4.1 Study Population and Global Database Utility

The present study analyzed a balanced longitudinal panel comprising 204 countries and territories over a 24-year period (2000–2023), yielding 4,896 country-year observations. This comprehensive global dataset provides a unique opportunity to examine long-term macroepidemiological trends in CVD mortality while accounting for temporal changes and country-specific heterogeneity. Compared with single-country cohort studies, the inclusion of nearly all WHO Member States substantially enhances the external validity and global representativeness of the findings. Similar ecological analyses based on the GBD database have demonstrated the value of multinational panel data for evaluating cardiovascular trends worldwide and informing international health policy (1–3).

Nevertheless, because the analyses were conducted using country-level aggregated estimates rather than individual patient data, the observed associations should be interpreted as population-level relationships. Consequently, causal inference at the individual level cannot be established, and the possibility of ecological fallacy should be acknowledged (4). Furthermore, national averages may conceal important within-country disparities related to socioeconomic status, ethnicity, urbanization, healthcare accessibility, and regional inequalities.

### 4.2 Baseline characteristics and global socioeconomic disparities

A marked socioeconomic gradient was observed across income groups, with low-income countries experiencing substantially higher age-standardized CVD mortality than high-income countries did (333.37 ± 148.13 vs. 208.44 ± 101.89 deaths per 100,000 people). This finding aligns closely with previous global analyses demonstrating that cardiovascular mortality disproportionately affects low- and middle-income countries (LMICs), where nearly three-quarters of all cardiovascular deaths occur (5,6).

One plausible explanation is the unequal distribution of healthcare resources. Hypertension screening programs, access to essential cardiovascular medications, preventive services, and long-term risk factor management remain considerably less available in resource-constrained settings (7,8). Consequently, elevated SBP contributed disproportionately to mortality in low-income countries (196.44 ± 92.35 deaths per 100,000), emphasizing persistent inequalities in primary cardiovascular prevention.

Geographically, several Pacific Island nations—including Nauru, Tuvalu, Kiribati, and the Marshall Islands—display exceptionally high CVD mortality throughout the study period. Previous reports have identified Oceania as one of the world’s highest-risk regions because of rapid nutritional transition, increasing obesity prevalence, widespread consumption of processed foods, and limited healthcare infrastructure (9,10). In contrast, countries such as Uzbekistan, Turkmenistan, and Ukraine have demonstrated declining mortality trends over time, suggesting that national cardiovascular prevention programs, improved hypertension management, and expanded healthcare access can substantially reduce cardiovascular mortality despite historically high baseline risk (11).

### 4.3 Temporal trends in CVD mortality and risk-attributable mortality

Global age-standardized CVD mortality declined steadily between 2000 and 2023 (∼340 to <260 deaths per 100,000 people), reflecting decades of progress in cardiovascular prevention, early diagnosis, improved pharmacological treatment, and acute cardiovascular care (5,12). These findings are consistent with previous GBD analyses demonstrating substantial reductions in age-standardized cardiovascular mortality despite continued increases in absolute numbers of deaths resulting from population growth and aging (1,5).

The temporary interruption of this decline from 2020–2021 likely reflects both direct and indirect effects of the COVID-19 pandemic. SARS-CoV-2 infection has been associated with myocardial injury, myocarditis, thromboembolic complications, arrhythmias, and acute coronary syndrome (13,14). Simultaneously, the disruption of healthcare systems, delayed emergency presentations, interruption of outpatient follow-up, and reduced access to chronic disease management have contributed to excess cardiovascular mortality worldwide (15).

The risk factor trajectories differed considerably. Smoking-attributable mortality and LDL cholesterol-attributable mortality declined substantially (37.35% and 30.00% reductions, respectively), likely reflecting successful tobacco control policies, the implementation of the WHO Framework Convention on Tobacco Control (FCTC), and the widespread use of statin therapy (16–18). Conversely, BMI-attributable mortality has increased modestly over time (+1.38%), which is consistent with the continuing global obesity epidemic and the increasing prevalence of metabolic syndrome (19,20).

### 4.4 Pairwise Correlations and Univariable Associations

Strong positive correlations were observed between overall CVD mortality and each major metabolic risk factor, particularly SBP (r = 0.99), LDL cholesterol (r = 0.92), fasting plasma glucose (r = 0.82), and BMI (r = 0.79). These findings are biologically plausible because hypertension, dyslipidemia, diabetes mellitus, and obesity frequently coexist within metabolic syndrome and collectively accelerate atherosclerosis and vascular injury (21,22).

However, these unadjusted associations should be interpreted cautiously because substantial collinearity exists among metabolic risk factors. Individuals and populations with obesity often simultaneously exhibit hypertension, dyslipidemia, insulin resistance, and chronic inflammation, making it difficult for univariable analyses to distinguish independent effects (23). Accordingly, multivariable longitudinal models are needed to estimate the unique contribution of each risk factor after accounting for confounding pathways.

### 4.5 Primary two-way fixed-effects (TWFE) analysis

After controlling for country-specific characteristics, secular trends, and socioeconomic development via the TWFE model with Driscoll–Kraay robust standard errors, elevated SBP emerged as the strongest independent predictor of CVD mortality (beta = 1.129, P < 0.001). This finding is consistent with extensive epidemiological evidence demonstrating that hypertension remains the leading modifiable determinant of cardiovascular mortality globally (24–26).

Elevated LDL cholesterol (beta = 0.622), smoking (beta = 0.452), and fasting plasma glucose (beta = 0.410) also retained statistically significant positive associations (P < 0.001 for all), which is consistent with decades of clinical and epidemiological evidence linking these factors to atherosclerosis, ischemic heart disease, and stroke (17,21,27).

In contrast, BMI was not independently associated with mortality after adjustment (beta = 0.049, P = 0.421). Rather than suggesting that obesity is clinically benign, this finding indicates that many of the cardiovascular effects of obesity are mediated through downstream metabolic abnormalities—including hypertension, insulin resistance, dyslipidemia, systemic inflammation, and endothelial dysfunction—which were simultaneously included in the multivariable model (20,28,29). Therefore, obesity remains an important upstream determinant of cardiovascular risk even when its direct statistical association is attenuated.

### 4.6 Sensitivity Analysis Comparing Country Fixed Effects and Two-Way Fixed Effects

Comparison of the country fixed effects model with the TWFE specification illustrates the importance of controlling for global secular trends. Under the country-only model, BMI demonstrated an apparent inverse association with CVD mortality (beta = −0.324), resembling the so-called "obesity paradox" reported in selected clinical populations (30).

However, after year fixed effects were introduced, this negative association disappeared (beta = +0.049). This finding strongly suggests that the paradox was attributable primarily to omitted variable bias rather than a true protective effect of obesity. During the study period, obesity prevalence increased globally, whereas age-standardized CVD mortality simultaneously declined because of improvements in prevention, treatment, and healthcare systems. Failure to adjust for common temporal trends can therefore produce misleading inverse associations between obesity and mortality (31).

### 4.7 COVID-19 exclusion sensitivity analysis

Excluding observations from 2020–2021 produced minimal changes in the estimated coefficients, indicating that the principal findings were robust despite the unprecedented disruption caused by the COVID-19 pandemic. SBP (beta = 1.123), LDL cholesterol (beta = 0.626), and smoking (beta = 0.447) remained significant predictors of CVD mortality.

Although fasting plasma glucose lost statistical significance after exclusion of the pandemic years (beta = 0.400, P = 0.125), the estimated coefficient remained remarkably stable. This suggests that the reduced significance likely reflects diminished statistical power after the removal of 408 observations rather than a meaningful biological change. Similar stability across sensitivity analyses strengthens confidence in the robustness of the primary findings (32).

### 4.8 Three-Year Lagged Analysis

The persistence of significant associations after introducing three-year exposure lags supports the temporal sequence between cardiometabolic risk factors and cardiovascular mortality. Hypertension, dyslipidemia, and tobacco exposure promote cumulative endothelial injury, vascular remodeling, and progressive atherosclerosis over many years before clinical cardiovascular events become evident (21,24). The particularly strong lagged association for LDL cholesterol (beta = 1.073, P < 0.001) agrees with evidence from lifelong exposure studies demonstrating that the cumulative LDL burden substantially influences future cardiovascular risk (33).

### 4.9 Assessment of Nonlinearity

The quadratic terms for all major risk factors were not statistically significant, suggesting predominantly linear relationships across the observed exposure range. These findings support previous meta-analyses demonstrating approximately log-linear or near-linear associations between major cardiovascular risk factors and vascular mortality over broad exposure ranges (24,34).

From a public health perspective, these results imply that incremental reductions in population risk factors are likely to produce proportional reductions in cardiovascular mortality without evidence of major threshold effects.

### 4.10 Heterogeneity by country income group

The significant interaction between income level and cardiometabolic risk factors demonstrated that socioeconomic development modified cardiovascular risk relationships (joint Wald statistic = 36.13, P = 0.00031). Smoking exhibited stronger associations in upper-middle-income countries (beta = +0.966, P = 0.039), whereas the relative contribution of LDL cholesterol varied across economic settings. These findings are consistent with the epidemiological transition theory, whereby countries undergoing rapid economic development experience shifts from infectious diseases toward chronic cardiometabolic diseases, accompanied by changing behavioral and metabolic risk profiles (35).

### 4.11 Heterogeneity by WHO Region

Regional analyses further demonstrated substantial geographic heterogeneity (joint Wald statistic = 47.02, P = 0.00058). Although elevated SBP remained the dominant predictor across all WHO regions (total coefficients ranging from 0.958 to 1.308), LDL cholesterol exhibited particularly strong associations within the African region (beta = +1.312, P < 0.001). These observations likely reflect ongoing epidemiological transitions, limited access to lipid-lowering therapies, and expanding urbanization throughout sub-Saharan Africa (36).

Similarly, smoking demonstrated stronger positive interaction associations in Africa (beta = +0.833), the Americas (beta = +1.536), and the Western Pacific (beta = +1.299), reflecting regional differences in tobacco consumption, cigarette composition, healthcare accessibility, and the implementation of tobacco control policies (16,36).

### 4.12 Influence and robustness analyses

Several country-year observations have exhibited high statistical leverage, particularly for small island developing states and countries with exceptionally high cardiovascular burdens. Importantly, leave-one-country-out (LOCO) analyses demonstrated minimal changes in regression coefficients, indicating that the principal findings were not driven by a small number of influential observations. This robustness enhances confidence in the stability and generalizability of the estimated associations.

### 4.13 Model Diagnostics and Robust Statistical Inference

Diagnostic testing demonstrated substantial serial correlation (Durbin–Watson statistic = 0.0989), heteroskedasticity (Breusch–Pagan LM statistic = 618.18, P < 0.001), and cross-sectional dependence, which are common features of multinational longitudinal datasets (37). Had conventional ordinary least squares or standard clustered standard errors been used, the statistical uncertainty would likely have been underestimated.

The use of Driscoll–Kraay robust covariance estimation therefore represents an important methodological strength, providing valid statistical inference despite violations of classical regression assumptions (38).

### 4.14 Overall Pattern of Findings

Overall, elevated SBP consistently emerged as the strongest independent predictor of global CVD mortality, followed by elevated LDL cholesterol, smoking, and fasting plasma glucose. Although BMI was not independently associated after adjustment, its contribution appears to operate predominantly through downstream metabolic pathways. These findings reinforce current international cardiovascular prevention guidelines emphasizing aggressive blood pressure control together with comprehensive management of dyslipidemia, diabetes, obesity, and tobacco use (24,39).

### 4.15 Study strengths and limitations

The principal strengths of this investigation include its global coverage of 204 countries and territories over 24 consecutive years (2000–2023; N = 4,896 country-year observations), its balanced panel structure, and the application of the TWFE regression framework. By simultaneously controlling for time-invariant country characteristics and common global calendar–year shocks, combined with Driscoll–Kraay robust covariance estimation and extensive sensitivity checks (1–5-year lags, pandemic exclusion, nonlinearity testing, and influence diagnostics), the analytical design provides a structurally stable evaluation of macrolevel cardiovascular trends. However, several methodological limitations require careful consideration. First, primary metrics rely on GBD 2023 modeled estimates, which depend on spatial smoothing and predictive covariates in data-sparse locations. Analyzing point estimates without full draw-level uncertainty propagation introduces classical errors-in-variables bias that may understate parameter variance. Second, while TWFE models eliminate unobserved time-invariant country heterogeneity, they cannot capture time-varying, nation-specific policy shifts, healthcare infrastructure reforms, or macrolevel policy feedback loops where mortality surges trigger rapid public health interventions.

Finally, because this analysis was conducted at the aggregated national level, the observed associations describe macropopulation relationships and cannot be extrapolated to individual-level clinical risk or biological etiology (ecological fallacy hazard). Furthermore, because GBD comparative risk assessment applies population-attributable fractions to total CVD deaths, structural mathematical coupling exists between risk-attributable and overall CVD mortality, particularly for high SBP (r = 0.99). Finally, strong collinearity among cardiometabolic risk pathways causes downstream mediators (SBP, lipids, fasting glucose) to absorb the direct statistical variance of upstream drivers, explaining the attenuation of the BMI coefficient in multivariable modeling (beta = 0.049, P = 0.421).

## 5. CONCLUSIONS

Using a comprehensive global longitudinal dataset covering 204 countries and territories between 2000 and 2023, this study demonstrated that age-standardized CVD mortality declined worldwide despite persistent regional and socioeconomic disparities. Elevated SBP emerged as the strongest independent predictor of cardiovascular mortality, followed by elevated LDL cholesterol, smoking, and fasting plasma glucose, whereas the effect of BMI was largely mediated through downstream metabolic pathways.

These findings support continued prioritization of hypertension control, lipid management, diabetes prevention, obesity prevention, and comprehensive tobacco control policies. Greater investment in equitable cardiovascular prevention programs—particularly within low- and middle-income countries—is essential to accelerate progress toward reducing the global burden of cardiovascular disease and achieving the Sustainable Development Goals related to noncommunicable diseases.

## Acknowledgments

The author expresses sincere appreciation to ReseMeds Academy for their technical consultation during the statistical data organization and manuscript preparation. Per the International Committee of Medical Journal Editors (ICMJE) and American Heart Association guidelines on writing assistance, the author acknowledges the technical use of automated assistive language technologies (ChatGPT and Claude) for Python code refactoring, reference formatting, and initial grammatical editing. The author assumes sole responsibility for the scientific content, data integrity, and final approval of this manuscript. The author also acknowledges the IHME and the World Bank for providing publicly accessible global health datasets.

## CRediT Authorship Contribution Statement

**Khalid Mohammed Al-Dhayani:** Conceptualization, Methodology, Data curation, Formal analysis, Validation, Investigation, Visualization, Writing – original draft, Writing – review & editing.

## Data availability

The original data used in this study are publicly available from the IHME GBD Results Tool (https://ghdx.healthdata.org/gbd-results-tool) and the World Bank Open Data repository (https://data.worldbank.org). The harmonized country-year analytical dataset generated during this study has been deposited in Zenodo and is publicly accessible at Al-Dhayani KM. Country-Year Analytical Dataset for Global Trends in Metabolic Risk Factors and Cardiovascular Disease Mortality, 2000–2023. Version 01. Zenodo; 2026. https://doi.org/10.5281/zenodo.21740644.

## Supplemental Material

Supplemental Appendix

Tables S1–S15

Figures S1–S6

