## Supplementary Materials for "Global Trends in Cardiovascular Mortality and Major Cardiometabolic Risk Factors, 2000–2023: A Longitudinal Ecological Analysis of 204 Countries and Territories"

### Supplementary Results

**S1. Distribution of Study Variables**

Kernel density estimation and boxplot analyses demonstrated right-skewed distributions for several study variables, particularly GDP per capita, high body mass index (BMI), and high systolic blood pressure (SBP), reflecting substantial between-country heterogeneity in socioeconomic development and cardiometabolic risk burden (Figures S2–S3). Despite the presence of several extreme observations, no evidence suggested data entry errors or implausible values, and all observations were retained because they represented valid country-level estimates. The large sample size and subsequent use of fixed-effects models with Driscoll–Kraay robust standard errors minimized the influence of departures from normality on statistical inference.

**S2. Country-Specific Temporal Trends**

Country-level trend analyses demonstrated considerable heterogeneity in the magnitude of changes in age-standardized CVD mortality during 2000–2023 (Table S1). Most countries experienced statistically significant declines in mortality over the study period, including Ukraine and Sudan, whereas temporal trends in the Marshall Islands and Egypt did not reach statistical significance. Analysis of absolute mortality changes further demonstrated that Kazakhstan, Mongolia, and Russia experienced the largest reductions in CVD mortality, while several countries, including the Solomon Islands, Dominican Republic, Egypt, and Guinea-Bissau, showed limited improvement or increasing mortality over time (Table S9).

**S3. Additional Temporal Trend Analyses**

Annual percentage change analyses demonstrated a synchronized interruption of the long-term declining trend during 2020–2021 across overall CVD mortality and all major cardiometabolic risk factors (Table S2). This transient increase was followed by renewed declines during 2022–2023, consistent with recovery after the acute phase of the COVID-19 pandemic. Regional analyses similarly demonstrated marked geographic variation in long-term mortality reductions, with the greatest absolute improvements observed in Europe and comparatively smaller improvements or increasing mortality in several lower-resource regions (Figure S6).

**S4. Multicollinearity Assessment**

Additional regression diagnostics confirmed substantial collinearity among cardiometabolic risk factors. Variance inflation factors were highest for high LDL cholesterol and high SBP, indicating strong intercorrelations between these exposures (Table S5). Nevertheless, the consistency of parameter estimates across multiple model specifications suggests that multicollinearity did not materially alter the principal study conclusions. The primary two-way fixed-effects specification was therefore retained as the prespecified analytical model.

**S5. Model-Based Decomposition of Global Mortality Decline**

Model-based decomposition analyses estimated the relative contribution of individual cardiometabolic risk factors to the predicted decline in global age-standardized CVD mortality during the study period (Tables S7–S8). Reductions in SBP were associated with the largest proportion of the modeled decline, followed by decreases in LDL cholesterol, smoking, and fasting plasma glucose. These analyses are intended to illustrate the relative importance of each modeled exposure within the fitted regression framework and should not be interpreted as direct causal attribution.

**S6. Extended Lagged Analyses**

Additional lagged analyses using 1-year and 5-year exposure intervals produced findings consistent with the primary 3-year lag model (Tables S15–S16). High SBP and high LDL cholesterol remained significantly associated with subsequent CVD mortality across all lag structures, whereas smoking demonstrated persistent but more variable associations. These findings support the temporal robustness of the observed relationships and reduce the likelihood that the primary results were driven by short-term fluctuations.

**S7. Detailed Subgroup Analyses**

Expanded interaction analyses demonstrated statistically significant heterogeneity across both World Bank income groups and WHO regions (Tables S12–S14). Although the magnitude of associations differed across subgroups, elevated SBP remained consistently associated with higher CVD mortality in every income category and geographic region examined. Regional variation was most evident for smoking and LDL cholesterol, suggesting that the relative importance of individual cardiometabolic risk factors differs across global settings.

**S8. Influence and Residual Analyses**

Cook's distance diagnostics identified a limited number of influential country-year observations, primarily involving small island developing states and several densely populated countries (Table S4). However, leave-one-country-out analyses demonstrated minimal changes in the principal regression coefficients, confirming that the primary findings were not driven by individual high-leverage observations. Residual analyses additionally identified countries with consistently lower- or higher-than-predicted mortality relative to their modeled cardiometabolic risk profiles (Tables S6, S10, and S11), suggesting the potential influence of unmeasured health system, environmental, or policy-related determinants.

**S9. Residual Diagnostics**

Residual diagnostic plots demonstrated acceptable overall model performance despite expected deviations from classical regression assumptions in this large international panel dataset (Figure S1). Quantile–quantile plots showed moderate departures from normality at the upper tails of the distribution, while residual-versus-fitted plots demonstrated relatively constant variance with only minor heteroskedasticity among countries with the highest mortality rates. Together with formal diagnostic testing, these findings support the use of Driscoll–Kraay robust covariance estimation for valid statistical inference in the presence of serial correlation, heteroskedasticity, and cross-sectional dependence.

### Supplementary Appendix Tables

**Table S1. Top 10 Countries by Mean CVD Mortality and Associated 24-Year Trends**

| **Location** | **Mean CVD Mortality** | **Annual Trend (Coef)** | **P-value** |
| --- | --- | --- | --- |
| Nauru | 892.48 | -7.26 | < 0.001 |
| Tuvalu | 725.54 | -2.52 | < 0.001 |
| Marshall Islands | 708.39 | -0.09 | 0.763 |
| Egypt | 699.37 | -1.85 | 0.129 |
| Sudan | 683.32 | -11.08 | < 0.001 |
| Afghanistan | 657.56 | -6.03 | < 0.001 |
| Morocco | 620.18 | -8.11 | < 0.001 |
| Uzbekistan | 612.83 | -6.42 | < 0.001 |
| Ukraine | 592.24 | -12.50 | < 0.001 |
| Turkmenistan | 586.99 | -8.49 | < 0.001 |

*Abbreviations: CVD, Cardiovascular Disease.*
*Notes: Mean CVD Mortality Rate represents average age-standardized deaths per 100,000 population over the 2000–2023 period. The Annual Trend Coefficient is derived from ordinary least squares (OLS) regression where Calendar Year is the independent variable. P-values < 0.05 are considered statistically significant.*

**Table S2. Annual Percentage Change (APC) in Global Average Cardiovascular Mortality Attributable to Major Metabolic Risk Factors and Smoking (2001–2023)**

| **Year** | **CVD Mortality (%)** | **High BMI (%)** | **High FPG (%)** | **High SBP (%)** | **High LDL (%)** | **Smoking (%)** |
| --- | --- | --- | --- | --- | --- | --- |
| **2001** | -1.35 | 0.03 | -0.75 | -1.22 | -1.73 | -1.84 |
| **2002** | -0.89 | 0.64 | -0.40 | -0.66 | -1.32 | -1.47 |
| **2003** | -1.28 | 0.11 | -0.56 | -1.12 | -1.55 | -1.74 |
| **2004** | -1.53 | -0.39 | -0.98 | -1.40 | -2.00 | -2.16 |
| **2005** | -1.23 | -0.03 | -0.68 | -1.16 | -1.65 | -1.85 |
| **2006** | -1.46 | -0.34 | -0.98 | -1.38 | -1.93 | -2.43 |
| **2007** | -1.73 | -0.35 | -1.17 | -1.58 | -2.23 | -2.65 |
| **2008** | -1.51 | -0.41 | -1.04 | -1.34 | -2.15 | -2.86 |
| **2009** | -1.56 | -0.30 | -1.07 | -1.37 | -2.32 | -2.84 |
| **2010** | -1.48 | -0.18 | -1.10 | -1.41 | -2.14 | -2.64 |
| **2011** | -1.43 | -0.23 | -1.09 | -1.33 | -2.06 | -2.65 |
| **2012** | -1.15 | -0.13 | -0.70 | -1.21 | -1.73 | -2.36 |
| **2013** | -1.74 | -0.55 | -1.38 | -1.81 | -2.49 | -2.87 |
| **2014** | -1.53 | -0.32 | -1.11 | -1.57 | -2.13 | -2.71 |
| **2015** | -0.26 | 0.68 | 0.40 | -0.25 | -0.39 | -1.57 |
| **2016** | -1.99 | -0.70 | -1.59 | -1.99 | -2.32 | -2.75 |
| **2017** | -1.60 | -0.38 | -0.94 | -1.63 | -1.76 | -2.43 |
| **2018** | -1.54 | -0.37 | -0.79 | -1.56 | -1.38 | -1.99 |
| **2019** | -1.72 | -0.21 | -1.34 | -1.69 | -1.80 | -2.33 |
| **2020** | -0.33 | 0.86 | 0.25 | -0.23 | -0.51 | -0.91 |
| **2021** | 1.43 | 3.31 | 2.17 | 1.61 | 1.73 | 1.68 |
| **2022** | -0.80 | 0.15 | -0.19 | -0.68 | -0.69 | -1.89 |
| **2023** | -0.69 | 0.54 | -0.05 | -0.55 | -0.76 | -0.93 |

*Abbreviations: APC, Annual Percentage Change; BMI, Body Mass Index; FPG, Fasting Plasma Glucose; SBP, Systolic Blood Pressure; LDL, Low-Density Lipoprotein Cholesterol.*
*Notes: APC values represent the year-over-year percentage change in age-standardized cardiovascular mortality rates attributable to each specific risk factor.*
*Interpretation: Negative values indicate a reduction in the mortality rate compared to the preceding year. Positive values (e.g., the notable spikes observed in 2020 and 2021 across all risk factors) indicate an increase in the mortality rate, likely reflecting the compounding impact of the COVID-19 pandemic on cardiovascular and metabolic health outcomes.*

**Table S3. Summary of Total Percentage Change in Global Attributable CVD Mortality (2000–2023)**

| **Risk Factor** | **Mean Value (2000)** | **Mean Value (2023)** | **Total % Change (2000–2023)** |
| --- | --- | --- | --- |
| High BMI | 32.78 | 33.23 | 1.38% |
| High Glucose | 30.78 | 26.43 | -14.12% |
| High SBP | 196.35 | 151.83 | -22.67% |
| High LDL | 66.62 | 46.63 | -30.00% |
| Smoking | 34.66 | 21.71 | -37.35% |

*Abbreviations: BMI, Body Mass Index; SBP, Systolic Blood Pressure; LDL, Low-Density Lipoprotein.*
*Notes: Mean values represent the global average of age-standardized CVD deaths per 100,000 population attributable to each specific risk factor.*

**Table S4. Influence Analysis and Identification of Potentially Influential Country-Year Observations**

| Location | Year | CVD mortality | High BMI | High glucose | High SBP | High LDL | Smoking | Cook's D |
| --- | --- | --- | --- | --- | --- | --- | --- | --- |
| Micronesia (United States) | 2000 | 621.73 | 87.15 | 87.36 | 304.55 | 122.64 | 99.54 | 0.009756 |
| Papua New Guinea | 2000 | 474.95 | 29.87 | 24.46 | 201.00 | 86.54 | 43.97 | 0.009746 |
| Bangladesh | 2007 | 488.21 | 12.46 | 29.22 | 232.47 | 56.10 | 48.57 | 0.009425 |
| Bangladesh | 2006 | 486.18 | 11.99 | 28.24 | 231.15 | 55.47 | 47.91 | 0.009421 |
| Bangladesh | 2008 | 484.16 | 12.81 | 29.77 | 231.28 | 56.01 | 48.83 | 0.008879 |
| Marshall Islands | 2000 | 715.65 | 98.88 | 139.90 | 396.08 | 126.90 | 66.11 | 0.008848 |
| Papua New Guinea | 2001 | 470.53 | 29.86 | 24.39 | 201.40 | 85.87 | 43.30 | 0.008828 |
| Bangladesh | 2009 | 486.34 | 13.45 | 30.75 | 234.67 | 57.19 | 49.89 | 0.008447 |
| Bangladesh | 2005 | 470.86 | 11.22 | 26.31 | 223.56 | 52.96 | 46.07 | 0.008422 |
| Marshall Islands | 2001 | 702.57 | 97.66 | 138.07 | 388.83 | 124.44 | 64.61 | 0.008227 |

**Abbreviations:** BMI, body mass index; CVD, cardiovascular disease; LDL, low-density lipoprotein; SBP, systolic blood pressure.

**Notes:** A total of 372 observations (7.60% of the analytic dataset) exceeded the prespecified influence threshold. The table presents the 10 observations with the largest Cook's distance values. Influence diagnostics were used to assess the sensitivity of model estimates to individual country-year observations rather than to automatically exclude observations.

**Table S5. Multivariable Linear Regression Coefficients (Metabolic and Behavioral Risk Factors Only)**

| **Variable** | **Coefficient (β)** | **Std. Error** | **P-value** | **95% CI** | **VIF** | **Std. β** |
| --- | --- | --- | --- | --- | --- | --- |
| Intercept | 26.02 | 0.60 | < 0.001 | 24.84 to 27.19 | N/A | N/A |
| High SBP | 1.35 | 0.01 | < 0.001 | 1.34 to 1.37 | 19.37 | 0.853 |
| High LDL | 0.45 | 0.02 | < 0.001 | 0.41 to 0.50 | 28.66 | 0.105 |
| Smoking | 0.46 | 0.03 | < 0.001 | 0.41 to 0.51 | 9.29 | 0.067 |
| High Glucose | 0.39 | 0.03 | < 0.001 | 0.33 to 0.45 | 13.13 | 0.053 |
| High BMI | -0.40 | 0.02 | < 0.001 | -0.45 to -0.36 | 11.25 | -0.068 |

*Abbreviations: SBP, Systolic Blood Pressure; LDL, Low-Density Lipoprotein; CI, Confidence Interval; VIF, Variance Inflation Factor; Std. β, Standardized Beta.*
*Notes: Adjusted R-squared = 0.982. P-values < 0.05 are considered statistically significant. This model isolates the metabolic and behavioral risk factors by excluding socioeconomic indicators.*

**Table S6. Top 15 Countries with Lower-than-Predicted Cardiovascular Disease Mortality Based on Mean Model Residuals**

| **Rank** | **Country** | **Mean Residual (Observed − Predicted)** |
| --- | --- | --- |
| 1 | Congo | −5.210647 × 10⁻¹⁴ |
| 2 | Guinea-Bissau | −4.592623 × 10⁻¹⁴ |
| 3 | Saudi Arabia | −4.540812 × 10⁻¹⁴ |
| 4 | Myanmar | −4.476049 × 10⁻¹⁴ |
| 5 | Albania | −4.370578 × 10⁻¹⁴ |
| 6 | Chad | −4.252154 × 10⁻¹⁴ |
| 7 | Ghana | −4.089321 × 10⁻¹⁴ |
| 8 | North Korea | −3.944992 × 10⁻¹⁴ |
| 9 | Russia | −3.904284 × 10⁻¹⁴ |
| 10 | Tuvalu | −3.863576 × 10⁻¹⁴ |
| 11 | Moldova | −3.748853 × 10⁻¹⁴ |
| 12 | China | −3.641532 × 10⁻¹⁴ |
| 13 | Greenland | −3.523108 × 10⁻¹⁴ |
| 14 | Yemen | −3.523108 × 10⁻¹⁴ |
| 15 | Bosnia and Herzegovina | −3.275158 × 10⁻¹⁴ |

**Abbreviation:** CVD, cardiovascular disease.

**Note:** Countries are ranked according to the mean residual (observed minus predicted CVD mortality) from the final multivariable regression model. Negative residual values indicate that the observed CVD mortality was lower than the model-predicted value. Residuals are presented in scientific notation exactly as generated by the statistical analysis.

**Table S7. Decomposition of Model-Predicted Decline in Global Age-Standardized CVD Mortality (2000–2023)**

| **Risk Factor Pathway** | **Absolute Contribution to Mortality Decline (deaths per 100,000)** | **Proportion of Predicted Decline (%)** |
| --- | --- | --- |
| **Systolic Blood Pressure (SBP)** | –50.24 | 70.91% |
| **LDL Cholesterol** | –12.44 | 17.55% |
| **Smoking** | –5.85 | 8.25% |
| **Fasting Plasma Glucose** | –1.78 | 2.52% |
| **Other Covariates (BMI, GDP, Urbanization)** | –0.55 | 0.77% |
| **2Total Model-Predicted Decline** | **–70.86** | **100.00%** |

**Note:** Mathematical decomposition of the global decline in age-standardized cardiovascular disease (CVD) mortality predicted by the primary Two-Way Fixed-Effects (TWFE) panel model. The absolute contribution of each risk factor was calculated by multiplying its adjusted regression coefficient ($\beta$) by the absolute global mean change in that specific risk-factor-attributable mortality metric between 2000 and 2023.

**Table S8. Decomposition of the Decline in Cardiovascular Disease Mortality Attributable to Major Risk Factors, 2000–2023**

| **Risk Factor** | **Absolute Change (2000–2023)** | **Estimated Contribution to CVD Mortality Change** | **Relative Contribution (%)** |
| --- | --- | --- | --- |
| High SBP | −44.52 | −50.24 | 70.91 |
| High LDL | −19.99 | −12.44 | 17.55 |
| Smoking | −12.95 | −5.85 | 8.25 |
| High glucose | −4.35 | −1.78 | 2.52 |
| Urban population | 7.21 | −0.37 | 0.52 |
| GDP per capita | 3814.38 | −0.20 | 0.28 |
| High BMI | 0.45 | 0.02 | −0.03 |

**Abbreviations:** BMI, body mass index; CVD, cardiovascular disease; GDP, gross domestic product; LDL, low-density lipoprotein cholesterol; SBP, systolic blood pressure.

**Note:** Absolute change represents the difference in the mean value of each risk factor between 2000 and 2023. Estimated contribution was calculated by multiplying the absolute change in each risk factor by its corresponding regression coefficient from the final multivariable model. Relative contribution (%) represents the proportional contribution of each risk factor to the modeled decline in CVD mortality over the study period.

**Table S9. Countries with the Largest and Smallest Changes in Age-Standardized Cardiovascular Disease Mortality, 2000–2023**

| **Panel A. Largest Reductions** |  |  |  |
| --- | --- | --- | --- |
|  | **Mortality 2000** | **Mortality 2023** | **Absolute Change** |
| Kazakhstan | 681.61 | 294.38 | −387.22 |
| Mongolia | 654.34 | 315.78 | −338.56 |
| Russia | 700.79 | 373.17 | −327.62 |
| Bahrain | 497.59 | 202.53 | −295.06 |
| Serbia | 703.55 | 421.42 | −282.14 |
| Estonia | 498.43 | 218.01 | −280.41 |
| Ukraine | 704.45 | 427.55 | −276.90 |
| Moldova | 592.42 | 316.74 | −275.68 |
| Georgia | 632.17 | 365.17 | −267.00 |
| Greenland | 505.65 | 250.70 | −254.95 |
| **Panel B. Smallest Reductions or Increases** |  |  |  |
| Senegal | 197.08 | 230.56 | +33.48 |
| Gambia | 204.22 | 239.48 | +35.26 |
| Burundi | 210.84 | 249.97 | +39.13 |
| Haiti | 539.82 | 584.42 | +44.60 |
| Tajikistan | 489.51 | 536.36 | +46.85 |
| Venezuela (Bolivarian Republic of) | 292.60 | 344.54 | +51.94 |
| Guinea-Bissau | 325.98 | 385.53 | +59.54 |
| Egypt | 640.59 | 703.82 | +63.23 |
| Dominican Republic | 218.86 | 308.05 | +89.20 |
| Solomon Islands | 553.24 | 686.88 | +133.64 |

**Abbreviation:** CVD, cardiovascular disease.

**Note:** Values represent age-standardized CVD mortality rates (deaths per 100,000 population). Absolute change was calculated as the difference between the 2023 and 2000 mortality rates. Negative values indicate reductions in mortality over the study period, whereas positive values indicate increases. Countries are ranked according to the magnitude of absolute change in CVD mortality between 2000 and 2023.

This format is suitable for submission to high-impact cardiovascular journals and is more compact than presenting two separate tables.

**Table S10. Top 10 Countries with Lower-than-Predicted Cardiovascular Disease Mortality Based on the Final Multivariable Model**

| **Rank** | **Country** | **Mean Residual (Observed − Predicted)** |
| --- | --- | --- |
| 1 | Moldova | −44.52 |
| 2 | Seychelles | −40.72 |
| 3 | Tajikistan | −40.15 |
| 4 | Bulgaria | −39.57 |
| 5 | Indonesia | −39.35 |
| 6 | Belarus | −38.72 |
| 7 | Kazakhstan | −36.38 |
| 8 | Georgia | −34.31 |
| 9 | Slovakia | −33.84 |
| 10 | Kyrgyzstan | −30.74 |

**Abbreviation:** CVD, cardiovascular disease.

**Note:** Countries are ranked according to the mean residual from the final multivariable regression model. Negative residuals indicate that observed CVD mortality was lower than predicted based on the modeled metabolic risk factors and socioeconomic covariates, suggesting better-than-expected performance.

**Table S11. Mean Model Residuals by World Bank Income Group**

| **Income Group** | **Mean Residual (Observed − Predicted)** |
| --- | --- |
| Upper-middle income | −12.18 |
| Lower-middle income | −1.77 |
| High income | −1.69 |
| Low income | 7.07 |

**Abbreviation:** CVD, cardiovascular disease.

**Note:** Mean residuals were calculated from the final multivariable regression model. Negative values indicate that observed CVD mortality was lower than predicted by the model, whereas positive values indicate higher-than-predicted mortality. Income groups were classified according to the World Bank.

**Table S12. Interaction Effects Between Metabolic Risk Factors and Income Groups or WHO Regions on Cardiovascular Disease Mortality**

| **Interaction Term** | **β Coefficient** | **SE** | **95% CI** | **P-value** | **Interaction Group** |
| --- | --- | --- | --- | --- | --- |
| High SBP × Low income | 0.150 | 0.205 | −0.251 to 0.552 | 0.463 | Income group |
| High LDL × Low income | −0.373 | 0.532 | −1.415 to 0.669 | 0.483 | Income group |
| High glucose × Low income | −0.277 | 0.785 | −1.817 to 1.262 | 0.724 | Income group |
| Smoking × Low income | 0.606 | 0.488 | −0.351 to 1.563 | 0.215 | Income group |
| High SBP × Africa | −0.077 | 0.120 | −0.312 to 0.158 | 0.522 | WHO region |
| High SBP × Americas | 0.273 | 0.164 | −0.049 to 0.595 | 0.097 | WHO region |
| High SBP × Eastern Mediterranean | 0.039 | 0.139 | −0.233 to 0.310 | 0.780 | WHO region |
| High SBP × South-East Asia | 0.152 | 0.200 | −0.239 to 0.543 | 0.446 | WHO region |
| High SBP × Western Pacific | 0.208 | 0.113 | −0.013 to 0.430 | 0.066 | WHO region |
| High LDL × Africa | 1.312 | 0.363 | 0.601 to 2.023 | <0.001 | WHO region |
| High LDL × Americas | 0.032 | 0.397 | −0.747 to 0.811 | 0.936 | WHO region |
| High LDL × Eastern Mediterranean | 0.426 | 0.396 | −0.350 to 1.203 | 0.282 | WHO region |
| High LDL × South-East Asia | 0.663 | 0.396 | −0.113 to 1.438 | 0.094 | WHO region |
| High LDL × Western Pacific | 0.615 | 0.300 | 0.027 to 1.203 | 0.041 | WHO region |
| High glucose × Africa | −0.718 | 0.611 | −1.916 to 0.480 | 0.240 | WHO region |
| High glucose × Americas | −0.562 | 0.771 | −2.074 to 0.950 | 0.467 | WHO region |
| High glucose × Eastern Mediterranean | −1.483 | 0.611 | −2.681 to −0.284 | 0.015 | WHO region |
| High glucose × South-East Asia | −0.398 | 0.774 | −1.915 to 1.119 | 0.607 | WHO region |
| High glucose × Western Pacific | −1.692 | 0.578 | −2.826 to −0.559 | 0.003 | WHO region |
| Smoking × Africa | −1.143 | 0.423 | −1.973 to −0.313 | 0.007 | WHO region |
| Smoking × Americas | −0.241 | 0.640 | −1.496 to 1.015 | 0.707 | WHO region |
| Smoking × Eastern Mediterranean | 0.058 | 0.546 | −1.013 to 1.130 | 0.915 | WHO region |
| Smoking × South-East Asia | −1.198 | 0.411 | −2.003 to −0.393 | 0.004 | WHO region |
| Smoking × Western Pacific | −0.596 | 0.341 | −1.265 to 0.073 | 0.081 | WHO region |

**Abbreviations:** β, regression coefficient; BMI, body mass index; CI, confidence interval; CVD, cardiovascular disease; LDL, low-density lipoprotein cholesterol; SBP, systolic blood pressure; SE, standard error.

**Note:** Interaction coefficients were estimated from the fully adjusted multivariable regression model to evaluate whether associations between metabolic risk factors and CVD mortality differed across World Bank income groups and WHO regions. Statistically significant interaction terms (P < 0.05). No significant interactions were observed for income group, whereas several significant regional interactions were identified, indicating geographic heterogeneity in the associations between metabolic risk factors and CVD mortality.

**Table S13. Total Subgroup-Specific Regression Coefficients by World Bank Income Classification**

| **Risk Factor** | **High Income (Reference)** | **Low Income** | **Lower-Middle Income** | **Upper-Middle Income** |
| --- | --- | --- | --- | --- |
| **High SBP** | 1.027 | 1.177 | 1.123 | 1.299 |
| **High LDL** | 0.829 | 0.456 | 0.689 | –0.793 |
| **High Fasting Glucose** | 0.990 | 0.713 | 0.215 | 1.526 |
| **Smoking** | –0.125 | 0.481 | 0.526 | 0.840 |

**Note:** Total subgroup-specific coefficients representing the adjusted population-level association between cardiometabolic risk factors and CVD mortality, stratified by World Bank income classification. Values represent the linear combination of the baseline coefficient (High Income reference group) and the income-specific interaction term. High systolic blood pressure (SBP) maintains a robust, uniformly positive association with CVD mortality across all economic strata. Conversely, structural heterogeneity is evident in other factors; for example, the association between smoking and CVD mortality is negligible in High-Income nations but strongly positive in Low- and Middle-Income nations, likely reflecting variations in smoking intensity, duration, and the availability of secondary cardiovascular interventions.

**Table S14. Total Subgroup-Specific Regression Coefficients by World Health Organization (WHO) Region**

| **Risk Factor** | **EURO (Reference)** | **AFRO** | **AMRO** | **EMRO** | **SEARO** | **WPRO** |
| --- | --- | --- | --- | --- | --- | --- |
| **High SBP** | 1.035 | 0.958 | 1.308 | 1.074 | 1.187 | 1.243 |
| **High LDL** | 0.146 | 1.458 | 0.178 | 0.573 | 0.809 | 0.761 |
| **High Fasting Glucose** | 1.262 | 0.544 | 0.701 | –0.220 | 0.864 | –0.430 |
| **Smoking** | 0.984 | –0.159 | 0.743 | 1.042 | –0.214 | 0.388 |

**Abbreviations**: AFRO, African Region; AMRO, Region of the Americas; EMRO, Eastern Mediterranean Region; EURO, European Region; SEARO, South-East Asia Region; WPRO, Western Pacific Region.

**Note:** Total subgroup-specific coefficients representing the adjusted population-level association between cardiometabolic risk factors and CVD mortality, stratified by WHO region. Values represent the linear combination of the baseline coefficient (European Region [EURO] reference group) and the region-specific interaction term. While High SBP remains a dominant factor globally (coefficients ranging from 0.958 to 1.308), profound geographic heterogeneity exists among other metabolic markers. High LDL cholesterol exhibits an exceptionally strong association with CVD mortality in the African Region (AFRO; 1.458) compared to EURO (0.146).

**Table S15. Sensitivity Analysis Using 1-Year and 5-Year Lagged Risk Factor Variables**

| **Variable** | **β (1-Year Lag)** | **P-value** | **β (5-Year Lag)** | **P-value** |
| --- | --- | --- | --- | --- |
| High SBP | 1.0589 | <0.001 | 0.7315 | <0.001 |
| High LDL | 0.7670 | <0.001 | 1.3579 | <0.001 |
| High Glucose | -0.0028 | 0.984 | -1.4153 | <0.001 |
| Smoking | 0.5949 | <0.001 | 0.5665 | 0.098 |
| High BMI | -0.0960 | 0.435 | -0.5826 | 0.016 |
| GDP per Capita | -0.0002 | <0.001 | -0.0004 | <0.001 |
| Urban Population (%) | 0.0943 | 0.200 | 0.7105 | <0.001 |

**Abbreviations:** SBP, systolic blood pressure; LDL, low-density lipoprotein cholesterol; BMI, body mass index; β, regression coefficient.

**Table note:** Results are from two-way fixed-effects panel regression models in which explanatory variables were lagged by 1 and 5 years relative to age-standardized cardiovascular disease (CVD) mortality. Regression coefficients (β) represent the estimated association between lagged risk factor–attributable mortality measures (or covariates) and CVD mortality after adjustment for country and year fixed effects. P-values <0.05 were considered statistically significant. This analysis was performed as a sensitivity analysis to evaluate the temporal robustness of the primary findings.

**Table S16. Sensitivity Analysis Using Alternative Lag Structures (1-Year and 5-Year Lags)**

| **Variable** | **β (1-Year Lag)** | **P-value** | **β (5-Year Lag)** | **P-value** |
| --- | --- | --- | --- | --- |
| High SBP | 1.0589 | <0.001 | 0.7315 | <0.001 |
| High LDL | 0.7670 | <0.001 | 1.3579 | <0.001 |
| High Glucose | −0.0028 | 0.984 | −1.4153 | <0.001 |
| Smoking | 0.5949 | <0.001 | 0.5665 | 0.098 |
| High BMI | −0.0960 | 0.435 | −0.5826 | 0.016 |
| GDP per Capita | −0.0002 | <0.001 | −0.0004 | <0.001 |
| Urban Population (%) | 0.0943 | 0.200 | 0.7105 | <0.001 |

**Abbreviations:** β, regression coefficient; SBP, systolic blood pressure; LDL, low-density lipoprotein cholesterol; BMI, body mass index.

**Note:** Results are from two-way fixed-effects panel regression models with country and year fixed effects. Explanatory variables were lagged by 1 year and 5 years relative to age-standardized cardiovascular disease (CVD) mortality. β coefficients represent the adjusted association between lagged GBD risk-attributable CVD mortality estimates (or covariates) and overall age-standardized CVD mortality. Significant associations were defined as *P* < 0.05. High SBP- and high LDL-attributable mortality remained significantly associated with CVD mortality across both lag structures, supporting the temporal robustness of the primary findings, whereas associations for high glucose, high BMI, smoking, and urbanization varied according to the lag period. GDP per capita remained inversely associated with CVD mortality in both models.

### Supplementary Appendix Figure

**Supplementary Figure 1. Model diagnostics for the primary linear mixed-effects regression model.**
**Legend:** Diagnostic plots evaluating the core statistical assumptions of the linear mixed-effects model **predicting** age-standardized cardiovascular disease mortality. **(A)** A scatter plot of model residuals versus fitted values, utilized to assess homoscedasticity (constant variance). The red dashed line indicates zero residual error. The residuals are generally distributed symmetrically around zero, though slight fanning at the highest fitted values indicates mild heteroscedasticity at the extreme upper bounds of mortality. **(B)** A Quantile-Quantile (Q-Q) plot assessing the normality of the model residuals. The sample quantiles closely track the theoretical normal distribution (solid red line) across the central mass of the data. The observed deviations at the extreme upper and lower tails (heavy-tailed distribution) are common in large, country-level epidemiological datasets and do not substantially compromise the robustness of the mixed-effects estimates.

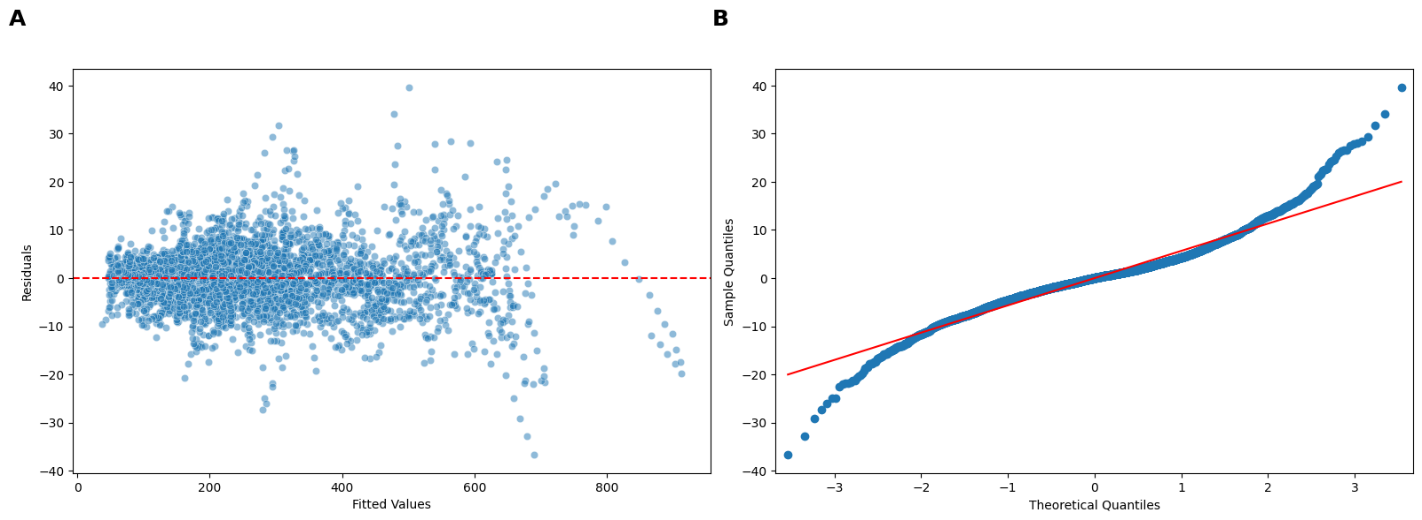

**Supplementary Figure 2. Pairwise scatter plot matrix and univariate distributions of primary study variables.**
**Legend:** A comprehensive scatter plot matrix visualizing the bivariate relationships and univariate distributions for age-standardized cardiovascular disease (CVD) mortality, key metabolic and behavioral risk factors (high systolic blood pressure [SBP], high LDL cholesterol, high fasting plasma glucose, and smoking), and macroeconomic status (GDP per capita). The diagonal panels display kernel density estimation (KDE) plots, illustrating the predominantly right-skewed distributions of the global health and economic metrics. The off-diagonal panels feature pairwise scatter plots overlaid with linear regression lines of best fit and 95% confidence intervals. The matrix visually confirms the strong positive collinearity among metabolic risk factors and their robust linear associations with overall CVD mortality. Furthermore, it highlights the pronounced inverse, non-linear (L-shaped) relationship between national wealth (GDP per capita) and both cardiovascular mortality and its underlying risk factors.

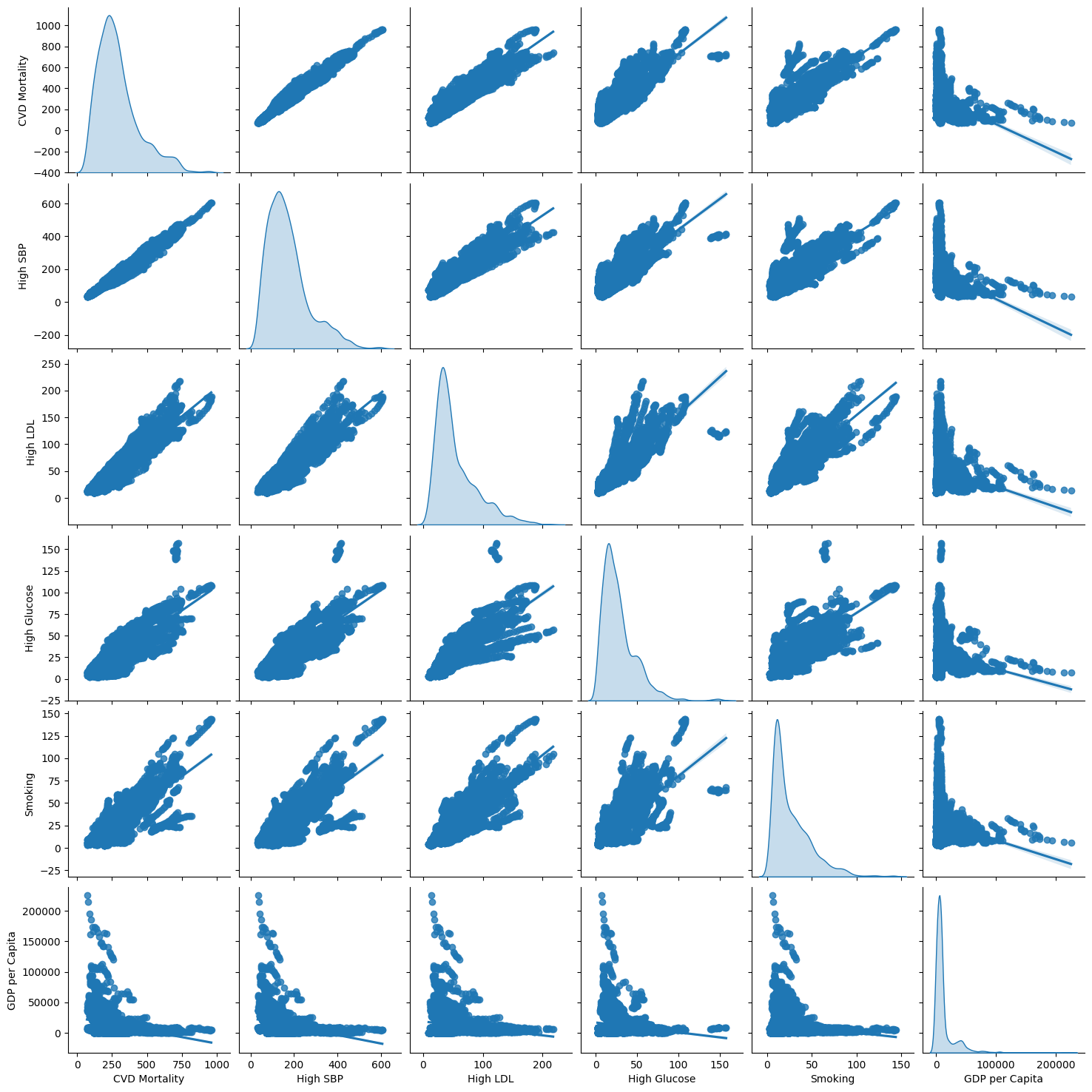

**Supplementary Figure 3. Distribution and outlier analysis of primary study variables across all country-year observations (2000–2023).**
**Legend:** Box-and-whisker plots displaying the statistical distribution of the primary outcome (CVD mortality), metabolic and behavioral risk factors, and socioeconomic covariates across the complete dataset of 4,896 country-year observations. The central horizontal line within each box represents the median value, while the upper and lower boundaries of the box indicate the interquartile range (IQR, 25th to 75th percentiles). The whiskers extend to 1.5 times the IQR, and individual circles represent outlier observations. The plots highlight the right-skewed nature and presence of extreme upper-bound outliers in several global health and economic metrics, particularly GDP per capita, high BMI, and high systolic blood pressure (SBP).

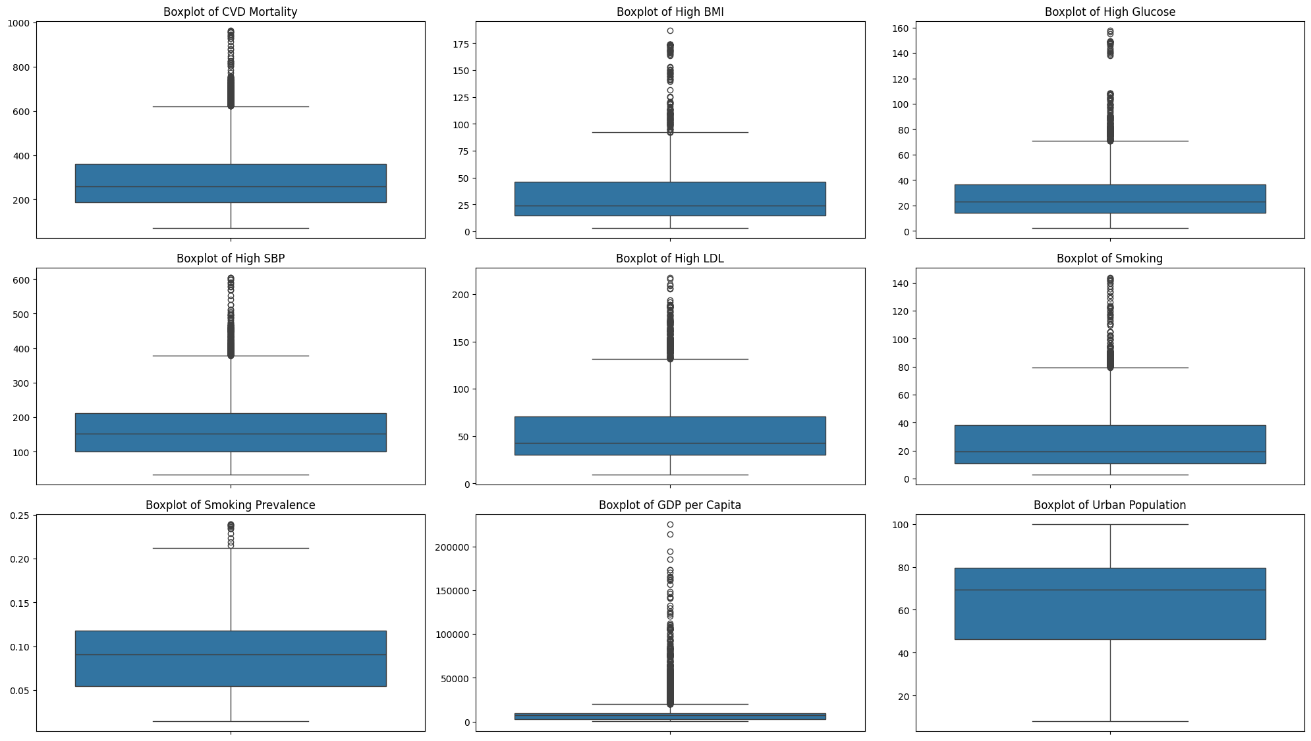

**Supplementary Figure 4. Longitudinal pathways of age-standardized cardiovascular disease (CVD) mortality rate for selected representative countries (2000–2023).**
*Legend:* Longitudinal trajectories of age-standardized CVD mortality rates per 100,000 population across four selected representative countries reflecting different epidemiological burdens: Nauru (purple line, very high burden), Tuvalu (blue line, high burden), Malawi (green line, moderate burden), and Peru (red line, low burden). While secular declines are visible for most countries over the 24-year period, a distinct, synchronized upward fluctuation is observed in 2021, coinciding with the acute phase of the COVID-19 pandemic, followed by a resumption of the downward trend in 2022–2023.

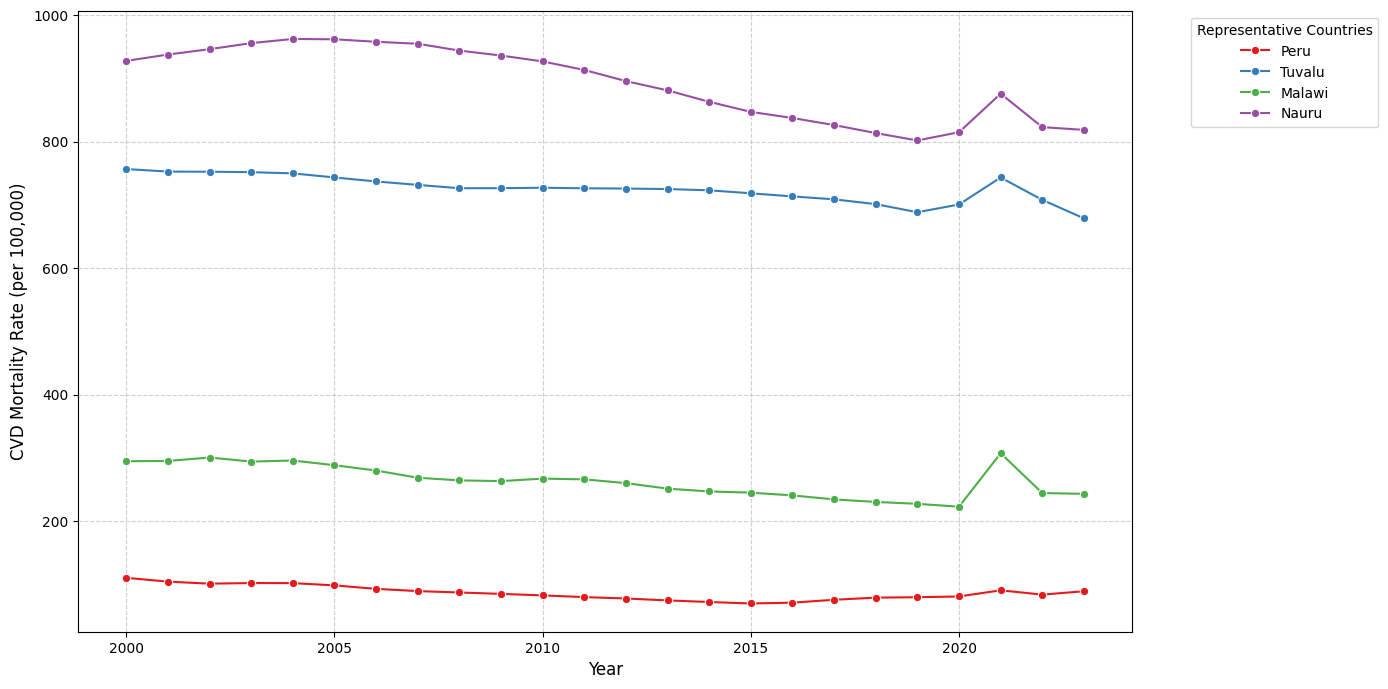

**Supplementary Figure 5. Influence analysis and outlier detection using Cook’s distance across all country-year observations.**

**Legend:** Diagnostic plot displaying the calculated Cook’s distance for each of the 4,896 country-year observations utilized in the primary multivariable regression model. The red dashed horizontal line signifies the standard influence threshold, calculated as 4/n (0.00082). While the vast majority of observations fall below this threshold, several distinct positive spikes denote highly influential data points. These prominent peaks predominantly represent specific statistical anomalies in small island developing states and densely populated nations (e.g., Micronesia, Papua New Guinea, Bangladesh, and the Marshall Islands). In total, 372 observations (7.60% of the dataset) exceeded the threshold. However, subsequent Leave-One-Country-Out (LOCO) sensitivity analyses confirmed that systematically excluding these highly influential countries did not substantially alter the core cardiometabolic coefficient estimates, verifying the structural robustness and stability of the global panel model.

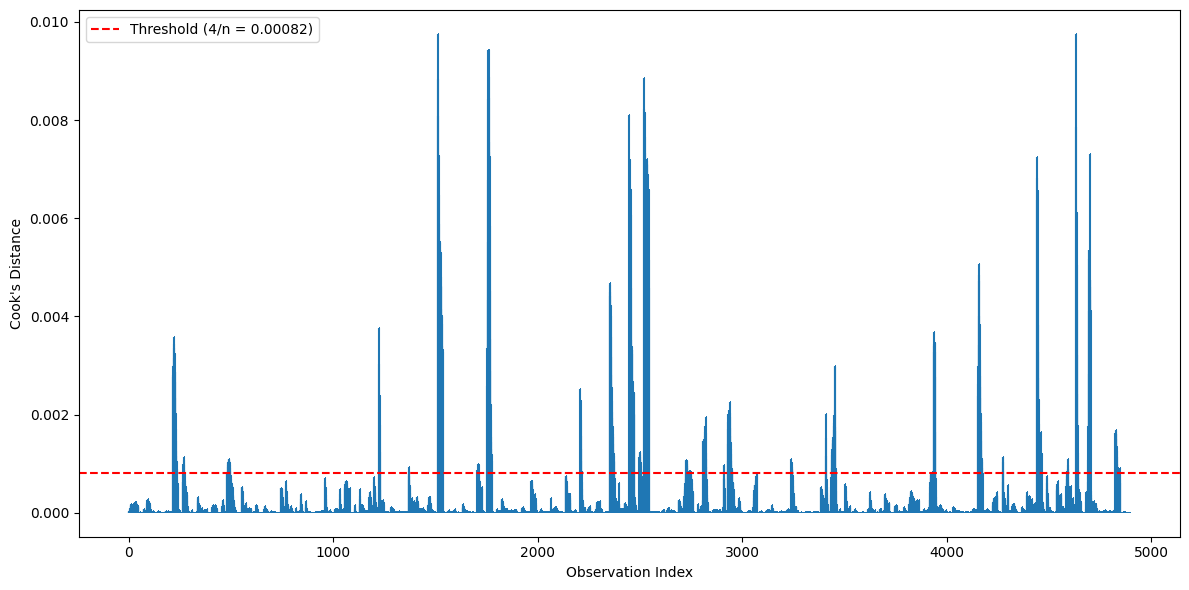

**Supplementary Figure 6. Average divergence in age-standardized cardiovascular disease (CVD) mortality rates by World Health Organization (WHO) region (2000–2023).**

**Legend:** Horizontal bar chart illustrating the mean absolute change in age-standardized CVD mortality rates (expressed as deaths per 100,000 population) across WHO regions over the 24-year study period. Negative values (purple/dark hues) indicate a net reduction in average mortality, whereas positive values (orange/red hues) indicate a net increase. The European Region (EURO) demonstrated the most substantial mean reduction in cardiovascular mortality (–283.79 deaths per 100,000), followed by the Eastern Mediterranean Region (EMRO; –115.92). In stark contrast, the Region of the Americas (AMRO) and the African Region (AFRO) experienced net average increases in CVD mortality (+44.60 and +59.54 deaths per 100,000, respectively). These regional divergences closely mirror broader socioeconomic gradients, wherein Upper-Middle and High-Income nations achieved massive absolute mortality reductions (–126.23 and –111.72, respectively), while Low-Income nations saw blunted progress (–38.91), highlighting severe and persisting structural disparities in global cardiovascular public health.

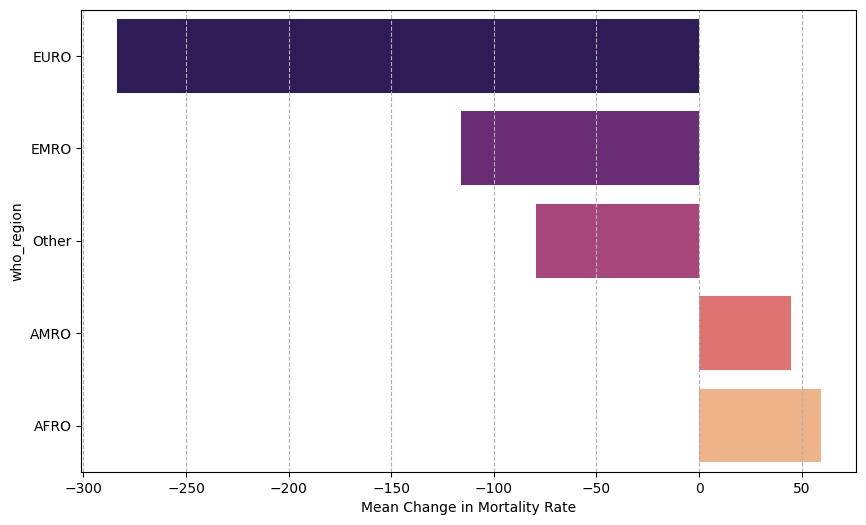
